# Long-Term Exposure to Fine Particulate Matter, Lung Function and Cognitive Performance: A Prospective Dutch Cohort Study on the Underlying Routes

**DOI:** 10.1101/2020.10.14.20212506

**Authors:** Benjamin Aretz, Fanny Janssen, Judith M. Vonk, Michael T. Heneka, H. Marike Boezen, Gabriele Doblhammer

## Abstract

**Background:** Exposure to fine particulate matter and black carbon is related to cognitive impairment and poor lung function, but less is known about the routes taken by different types of air pollutants to affect cognition.

**Objectives:** We tested two possible routes of fine particulate matter (PM_2.5_) and black carbon (BC) in impairing cognition, and evaluated their importance: a direct route over the olfactory nerve or the blood stream, and an indirect route over the lung.

**Methods:** We used longitudinal observational data for 31232 people aged 18+ from 2006 to 2015 from the Dutch Lifelines cohort study. By linking current and past home addresses to air pollution exposure data from ELAPSE, long-term average exposure (≥ ten years) to PM_2.5_ and BC was calculated. Lung function was assessed by spirometry and Global Initiative (GLI) z-scores of forced expiratory volume in 1s (FEV_1_) and forced vital capacity (FVC) were calculated. Cognitive performance was measured by cognitive processing time (CPT) assessed by the Cogstate Brief Battery. Linear structural equation modeling was performed to test the direct/indirect associations.

**Results:** Higher exposure to PM_2.5_ but not BC was directly related to higher CPT and thus slower cognitive processing speed [18.33 (×10^−3^) SD above the mean (95% CI: 6.84, 29.81)]. The direct association of PM_2.5_ constituted more than 97% of the total effect. Mediation by lung function was low for PM_2.5_ with a mediated proportion of 1.78% (FEV_1_) and 2.62% (FVC), but higher for BC (28.49% and 46.22% respectively).

**Discussion:** Our results emphasize the importance of the lung acting as a mediator in the relationship between both exposure to PM_2.5_ and BC, and cognitive performance. However, higher exposure to PM_2.5_ was mainly directly associated with worse cognitive performance, which emphasizes the health-relevance of fine particles due to their ability to reach vital organs directly.

## INTRODUCTION

Air pollution contributes substantially to the global burden of disease; it is responsible for 4.2 million deaths, about 8000 deaths per year in Europe (Lelieveld et al. 2019), and for 103.1 million lost years of healthy life globally in 2015 (Cohen et al. 2015).

There is also recent evidence that exposure to air pollution is associated with lower cognitive performance (Zhang et al. 2018) and a higher incidence of dementia (Carey et al. 2018). A cohort study from Germany investigated the relationship between air pollution and cognitive functioning as well as local brain atrophy measured by magnetic resonance imaging (Nußbaum et al. 2020). In this study, higher exposure to PM_2.5_ and PM_2.5 absorbance_ was related to lower cognitive functioning, and higher PM_2.5_ was additionally associated with local brain atrophy. A double-cross over experiment suggested that short-term exposure to PM_2.5_ had adverse effects on cognitive functioning measured by the Mini-Mental State Examination (MMSE) (Shehab and Pope 2019). A prospective cohort study among older Chinese adults, which also measured cognitive performance by using the MMSE, found that each 10-μg/m^3^ increase in PM_2.5_ was associated with a 5.1% increase in the risk of poor cognitive functioning (Wang et al. 2020). In the US, a doubling in BC level was related to 1.57 times higher odds of low MMSE scores (Colcino et al. 2017). A study, which explored the relationship between exposure to different constituent of PM_2.5_ and cognitive functioning among older Puerto Rican in the US, found that long-term exposure to BC, nickel, sulfur, silicon, and PM_2.5_ in general were related to slower mental speed or decreased recognition (Wurth et al. 2018).

Although, we know more about the negative effects of particulate matter on cognitive performance, the exact routes by which air pollutants may unfold their neurotoxic effects have barely been tested empirically, and there are substantial gaps in the knowledge of the underlying causal mechanisms (Griffiths and Mudway 2018). Commonly, two hypotheses are discussed (Block and Calderón-Garcidueñas 2009).

First, air pollutants may damage the brain directly by entering through the olfactory nerve or the lung, with subsequent entry into the blood stream providing access to the brain (**Figure 1**, path 1). It is mainly very fine particles which are assumed to follow this route (Block and Calderón-Garcidueñas 2009; González-Maciel et al. 2017). Second, air pollutants may enter the lung by inhalation, thus impairing lung function or causing pulmonary inflammation (**Figure 1**, path 2). After inhalation, especially fine particles can penetrate the deepest parts of the lung, e.g. the alveoli, due to their small size (Sturm 2020; Xing et al. 2020). Impaired lung function may cause lower (abnormal) blood oxygen levels (hypoxemia) leading to systemic inflammation, oxidative stress, cerebral arterial stiffness and small-vessel damage (Lutsey et al. 2018). Air pollutants also cause inflammatory responses of immune cells residing in the lung, e.g. pulmonary macrophages, thereby adding to or causing a substantial systemic presence of inflammatory mediators (Guarnieri and Balmes 2014).

**Figure 1.**
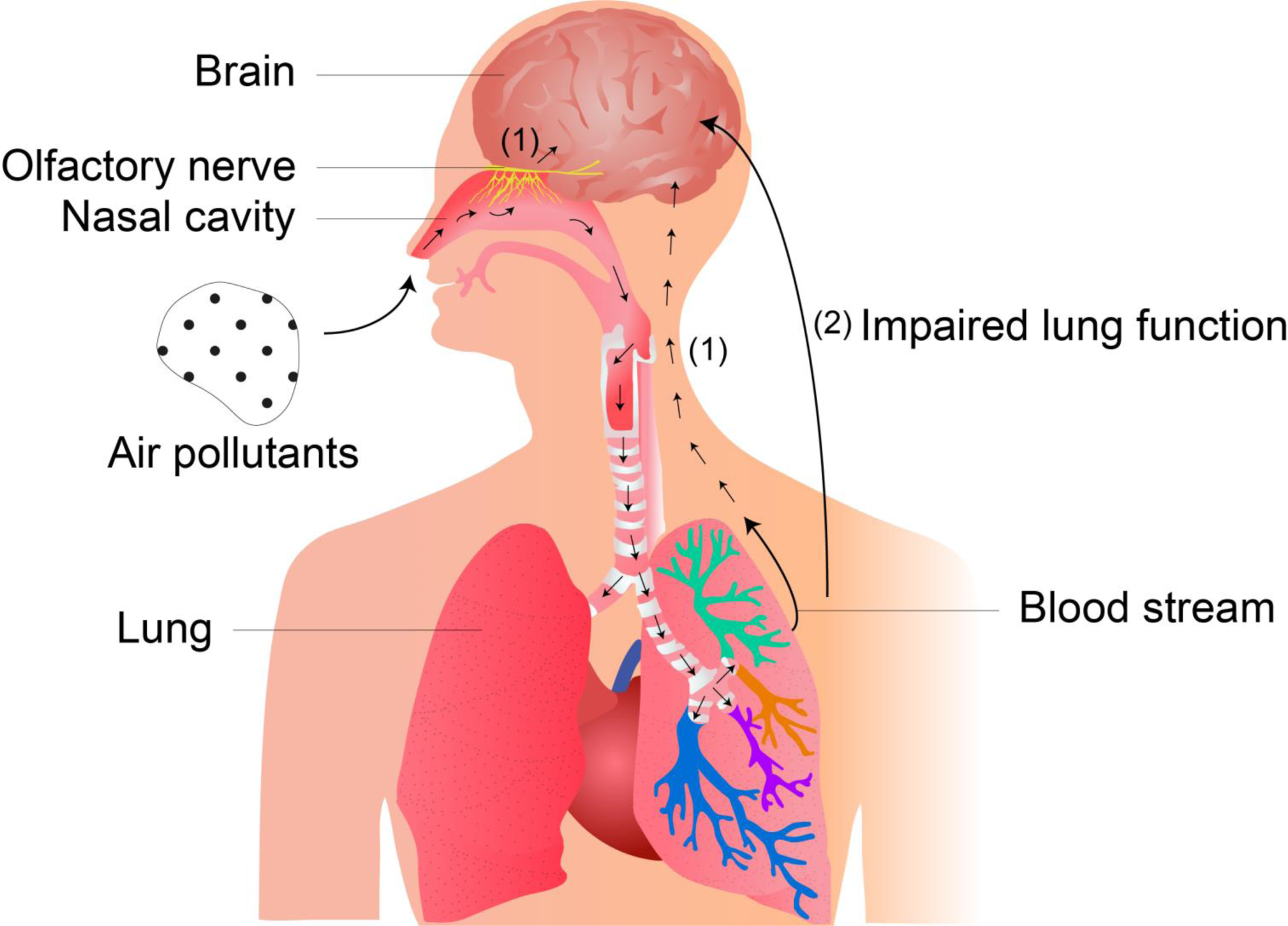
Routes Taken by Inhaled Fine Particles to Cause Subsequent Cognitive Impairments (source: authors’ drawing) Mainly smaller air pollutants (fine particles) may damage the brain directly by entering through the olfactory nerve or the lung, with subsequent entry into the blood stream providing access to the brain (**path 1**). Air pollutants may also enter the lung by inhalation, thus impairing lung function or causing pulmonary inflammation (**path 2**). Impaired lung function may cause lower (abnormal) blood oxygen levels (hypoxemia) leading to systemic inflammation, oxidative stress, cerebral arterial stiffness and small-vessel damage. Air pollutants also cause inflammatory responses of immune cells residing in the lung, e.g. pulmonary macrophages, thereby adding to or causing a substantial systemic presence of inflammatory mediators.

In accordance with this second hypothesis, previous research showed that ambient air pollution is a major health risk contributing substantially to respiratory mortality (Lelieveld et al. 2019), and air pollution is related to lower pulmonary function (Adam et al. 2015), higher COPD prevalence (Bloemsma et al. 2016), higher asthma prevalence (Zheng et al. 2015), higher lung cancer mortality (Dimakopoulou et al. 2014), and a higher burden of COPD (Cohen et al. 2015). But, the negative effect of small particulate matter on lung function, however, is still contested. A meta-analysis of five cohorts in the European Study of Cohorts for Air Pollution Effects (ESCAPE) observed that higher levels of NO_2_, but not of PM_10_ and PM_2.5_, were associated with lower levels of forced expiratory flow in one second (FEV_1_) and forced expiratory flow (FVC) among adults (Adam et al. 2015). On the contrary, the Framingham Heart Study found that also relatively low levels of PM_2.5_ were related to lower FEV1 and FVC, and an accelerated decline in lung function (Rice et al. 2015). For Black Carbon (BC), there is evidence from a women’s cohort from Boston, Massachusetts, that higher levels of BC were associated with decreased lung function in terms of FEV_1_ and FVC (Suglia et al. 2008).

Although the hypothesized direct and indirect pathways taken by inhaled air pollutants, there are hardly any cohort studies which explored the interrelations between air pollution, lung function and cognition. To our knowledge, only one cohort study exists that explored the mediating role of FEV_1_ and FVC in cognitive impairment caused by long-term exposure to NO_2_, PM_10_, and PM_2.5_ (Hüls et al. 2018). However, this study observed just a small, female cohort in Germany and did not find any significant mediation by lung function. But it showed that there was a total effect/ a general relationship in the way that higher exposure to fine particulate matter was related to poor cognitive performance.

Due to the existing lack of research in exploring the interrelation between air pollution, lung function and cognition, this new cohort study has thus two objectives. First, it empirically tests the hypothesized routes taken by fine particulate matter in general (PM_2.5_) and Black Carbon (BC), which is, next to the ultrafine particles (UFP), a major component of PM_2.5_., to impair people’s cognitive performance. Second, it evaluates the importance of the found routes in impairing cognition.

## METHODS

### Study Population and Design

We explored data from the Netherlands, a low pollution setting, and used longitudinal observational data for people aged 18+ from 2006 to 2015 from the Dutch Lifelines cohort study. Lifelines is a multi-generational prospective cohort study on multifactor risks for diseases, which recruited about 10% of the population in the three provinces of the Northern Netherlands, from whom 110,908 adults had a baseline and a follow-up assessment (Scholtens et al. 2015). Current and past residential addresses of each participant were obtained from municipal administration data. The Lifelines Cohort Study is conducted according to the principles of the Declaration of Helsinki and is approved by the medical ethical committee of the University Medical Center Groningen, The Netherlands.

In our study design, we tackled the issue of correct causal time order between the cause (exposure to fine particulate matter), mediator (lung function) and outcome (cognitive performance) of interest. For this purpose, we distinguished between three time periods. The first time period is the time up to baseline for which the air pollution exposure data over a minimum of ten years through the 31^st^ of December of the year before baseline were calculated for each participant. The second time period is the baseline (2006-2012), at which the participants were recruited for the Lifelines cohort study and the lung function was measured. And the third time period is the follow-up (2014-2015), at which the participants’ cognitive performance was assessed.

All participants aged 18+ with data available on residential addresses over a minimum of ten years through the 31^st^ of December of the year before baseline, with valid air pollution exposure data, a valid lung function measurement at baseline (2006-2012), and a valid measurement of cognitive performance at follow-up (2014-2015) were included (**Supplementary Figure S1)**. Our final sample size was 31232 people (see **Supplementary Table S3** for a comparison of the descriptive statistics among participants with complete and incomplete data).

### Outcome Assessment: Cognitive Performance Measured by Cognitive Processing Time (CPT)

To measure the participants’ cognitive performance in our follow-up period (2014-2015), we used the Cogstate Brief Battery (CBB), which is an age-specific validated standardized computerized tool to measure four domains of cognitive performance: psychomotor speed, visual attention, visual learning, and working memory (Lim et al. 2013). Other studies have used CBB to detect (mild) cognitive impairment (Maruff et al. 2009).

The CBB in Lifelines comprises four card tasks in the space of 11 minutes, reflecting the four cognitive domains: detection (2 min), identification (2 min), visual learning and memory (5 min), and working memory (2 min). Each task displays a textual instruction screen with a description of the task requirements. Each consists of several exercises (e.g. related to speed or accuracy) to be solved by selecting the “Yes” or “No” buttons on the screen (Lim et al. 2013).

Clinical practice has shown that the cognitive processing speed, defined as the ability to process information rapidly, is closely associated with the ability to solve (complex) cognitive tasks (Lichtenberger et al. 2013) and is thus one of the most important domains of cognitive performance (Salthouse and Ferrer-Caja 2003). Accordingly, we obtained a speed composite score using the speed measures from detection, identification and working memory task as primary outcomes in consultation with the Cogstate research team.

Each speed measure reflects the mean time for correct responses in each domain and was log (10)-transformed for better normality. For each of the three speed measures, we calculated z-standardized scores for the age groups: 18-34, 35-49, 50-59, 60-69, 70-79, 80-90, 90-99. A composite score measuring the overall cognitive processing time (CPT) was then computed by averaging the z-standardized speed scores of the three tasks. The final composite score was then z-standardized again. Positive scores mean that people had higher CPT (slower speed) and thus worse cognitive performance, and negative scores that they had lower CPT (faster speed) and thus better cognitive performance when compared to the mean. To control our cognitive speed outcome (CPT) for the accuracy of responses given, we accounted for age-standardized proportion of correct trials and the total number of trials per participant in the three CBB domains we used.

### Exposure Assessment: Fine Particulate Matter and Black Carbon

We used exposure data on two ambient air pollutants available in the Lifelines dataset: particulate matter (PM) of particulates with diameters of 2.5 µm and smaller (PM_2.5_) and the black carbon (BC) proportion in the PM_2.5_. The exposures were estimated by Lifelines using air pollution models developed in the project “Effects of Low-Level Air Pollution: A Study in Europe” (ELAPSE) (see de Hoogh et al. 2018 for a detailed description). In brief, satellite-derived and chemical transport model estimates were used to develop fine spatial scale land use regression (LUR) models for Western Europe for 2010. PM_2.5_ concentrations were derived from the European Air Quality Database (AirBase v8) and BC annual means from the monitoring campaign conducted in the “European Study of Cohorts for Air Pollution Effects” (ESCAPE) (Eeftens et al. 2012). The developed LUR models in ELAPSE for all included sites explained 62% of the variance PM_2.5_, whereas for BC 54% was explained.

To assess an individual’s long-term exposure to PM_2.5_ and BC up to our baseline period, we linked the current and past home addresses of the individuals to the average annual concentrations for PM_2.5_ and BC for that address for 2010 coming from the LUR developed in ELAPSE. For this purpose, we/lifelines used the Municipal Personal Record Database (Zijlema et al. 2016) that contains the home addresses and thus the geolocations of all individuals who live or have lived in the Netherlands so that residential mobility and length of exposure can be traced. For each air pollutant and individual, we calculated time-weighted average (*TWA*) concentrations by weighting the average exposure concentrations 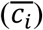 of a residential location in 2010 with the duration of residence at a specific location (exposure time of the data) (*t*_*i*_) (Cohen et al. 1996).

### Mediator Assessment: Lung Function

Lung function at baseline (2006-2012) was assessed by Lifelines through spirometry, performed by trained medical staff according to American Thoracic Society guidelines using a Welch Allyn Version 1.6.0.489 PC-based SpiroPerfect with CardioPerfect workstation software. We used two volume measures, the forced expiratory volume in one second (FEV_1_) and the forced vital capacity (FVC), as outcome variables. To come up with interpretable scores, we used age-, sex-and height-specific reference values for FEV_1_ and FVC as provided by the Global Lung Initiative (GLI) (Quanjer et al. 2012). Subsequently, the predicted proportion of the empirical lung function scores compared to the predicted reference scores were calculated for all both lung function measures. Scores beyond 100% mean that people had better lung parameters compared to the reference values, whereas scores lower than 100% mean that they had worse.

### Assessment of Potential Confounders

We controlled for age, sex, socio-demographic and lifestyle confounders, and respiratory and cognition-related diseases at baseline (2006-2012). Education level was defined as the highest education level completed (none, primary, lower secondary vocational, secondary vocational, senior general secondary, higher vocational, university, other). Income was measured by an individual’s net income per month (less than 1500 Euro, between 1500 and 2500 Euro, higher than 2500 Euro, unknown/ unexpressed). Body mass index (BMI) was calculated as the individual’s measured weight divided by measured height square and categorized in one of three groups: less than 25, between 25 and less than 30, and equal to or higher than 30. Hypertension was operationalized by a systolic pressure higher than 139 mmHg or a diastolic pressure higher than 89 mmHg. Systolic and diastolic blood pressure was measured by medical staff. Prevalence of respiratory diseases, namely asthma and COPD, as well as cognition-related diseases, namely multiple sclerosis, depression, diabetes (type I or II), and stroke, was derived by a question whether a doctor has ever diagnosed that specific disease in participants before (yes/ no). Pack-years of cigarettes smoked (1 pack-year = 20 cigarettes per day/ 1 year) were calculated from the baseline questionnaire collecting data on a person’s smoking history. Additionally, we controlled for the province of residence at baseline (Drenthe, Groningen, Friesland, other) and the total time period to which the long-term exposure to air pollution applied.

### Statistical Analyses

We used mediation analysis to explore whether the association between air pollution (X) and CPT (Y) followed a direct route to the brain or was mediated by lung function (*M*). We performed linear structural equation models (SEM) without feedback loops and with robust standard errors by Huber/ White (Breusch-Pagan-Test, p < 0.001). Exposure and confounders were treated as exogenous variables, and lung function as well as CPT were seen as endogenous variables. We introduced an outcome equation (equation 1) and a mediator equation (equation 2), whereby *C*_*n*_ denote the specific confounding variables, and *θ*_0_ to *θ*_*n*_, and *β*_0_ to *β*_*n*_ are the unobserved parameters (see also **Supplementary Figure S2** for the model approach):

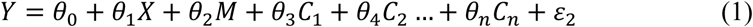

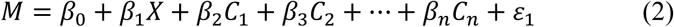

We estimated the two equations simultaneously and performed subsequently a decomposition of the total effect (the following usage of the words total, indirect or total effect does not imply causality; terms represent standard technical terms) *TE* = *β*_1_ + *θ*_1_ + *θ*_2_ into a direct effect *DE* = *θ*_1_ and the two separate indirect paths (*β*_1_, *θ*_2_). The both indirect path coefficients can be combined into one indirect effect *IE* = *β*_1_ × *θ*_2_ (Gunzler et al. 2013). To test the significance of the indirect effect (*IE*) we used the delta method/ Sobel Test (Sobel 1982). We calculated direct effect proportions 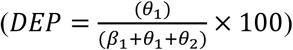 and indirect effect proportions 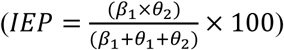 to quantify the importance of the significant direct effect and indirect effect compared to the total effect. The significance threshold was .05 and all tests were 2-sided. The calculations were performed using Stata MP 13.1.

We performed three kinds of sensitivity analyses. First, we checked the robustness of our SEM approach to ensure that our results were valid. We applied causal mediation analysis using a potential outcome approach with bootstrapping (1,000 iterations) for inference evaluation. Causal mediation analysis was performed by using the “mediation” package in Stata, which is based on the “mediation” package for causal mediation analysis in R (Tingley et al. 2014).

Second, we estimated SEMs for study participants aged 45 because the air pollution effects on cognitive performance are assumed especially evident for those ages when cognitive decline has generally started (Singh-Manoux et al. 2012).

Third, we used the average air pollution concentrations from ELAPSE at the baseline residential address only to check whether our sample selection confined to participants with historical addresses of at least ten years available biased the results.

## RESULTS

### Characteristics of the Study Participants

Of the 31232 participants 16103 (51.56%) had faster Cognitive Processing Time (CPT) and 15129 (48.44%) slower CPT than the mean at follow-up (2014-2015) (**Table 1**). The average daily exposure to PM_2.5_ was 14.95 [min = 9.58, max = 20.12] µg/m^3^, and 1.25 [min = 0.65, max = 2.34] µg/m^3^ to BC (**Figure 2**). The average time for which migration history and the linked air pollution data from ELAPSE at historical home address was available was 8567.56 days (23.46 years). For lung function, the average FEV_1_ was 3.55 liters (96.62% predicted) and FVC 4.60 liters (100.69% predicted) (**Table 1**).

**Table 1.**
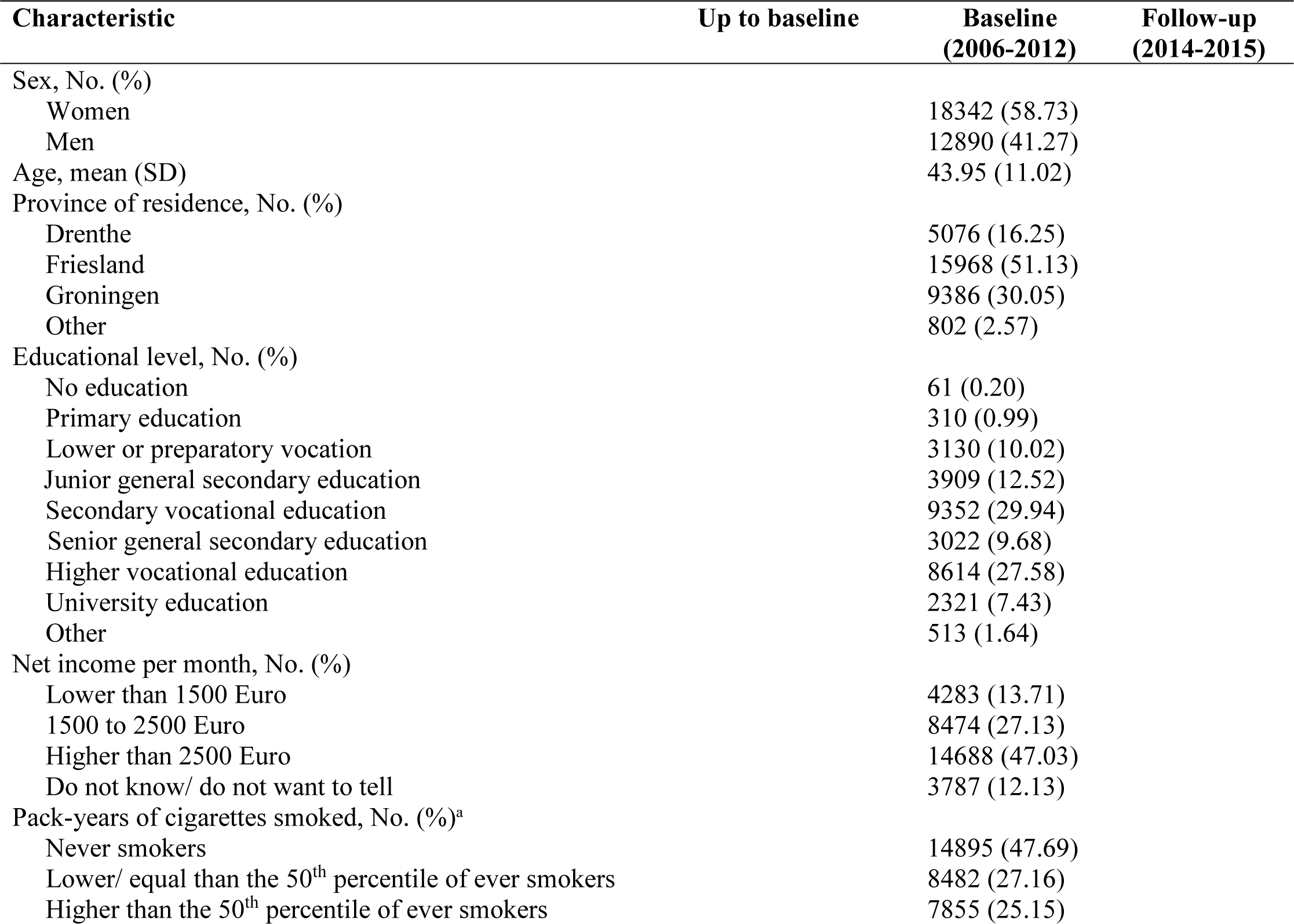
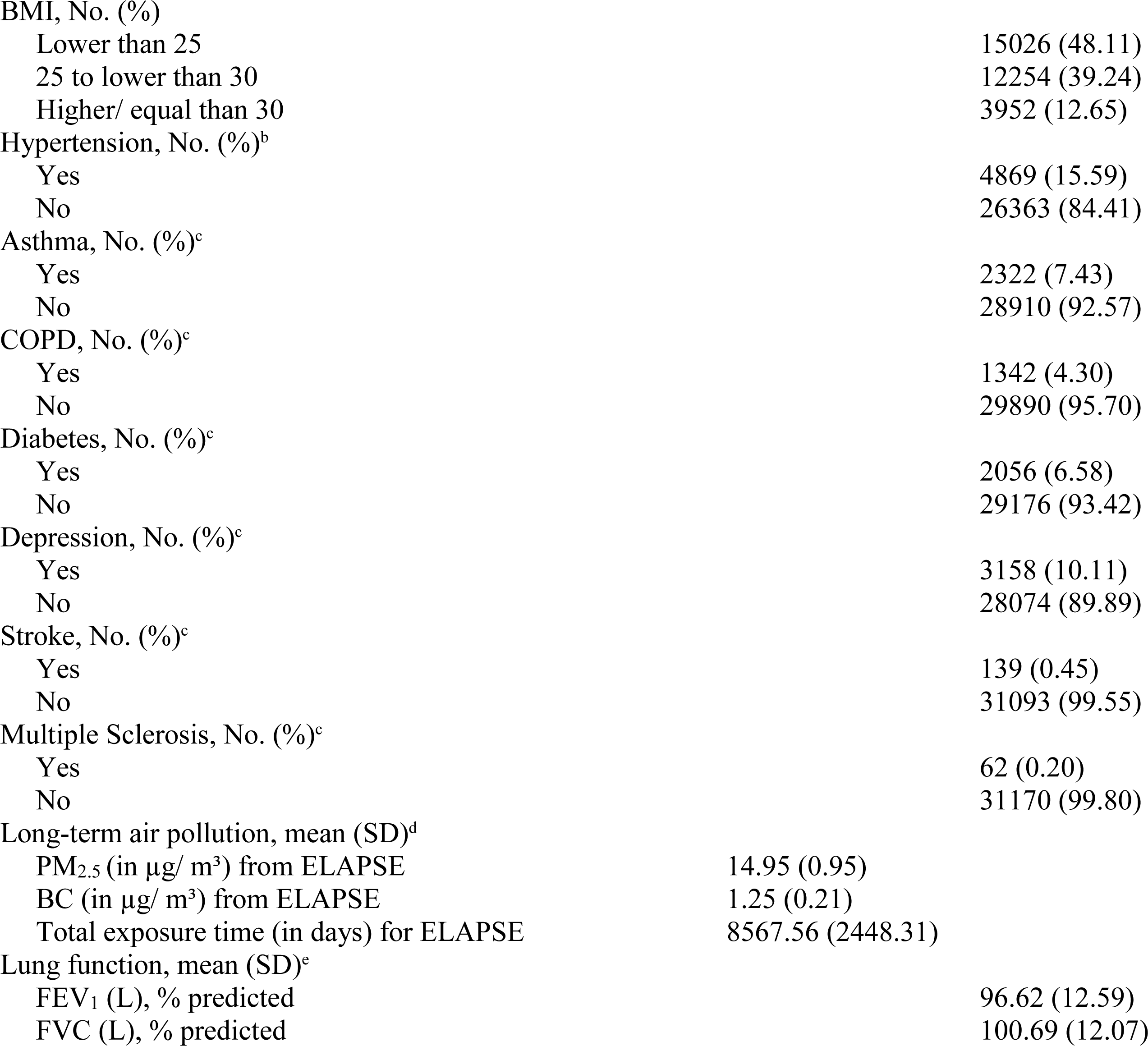
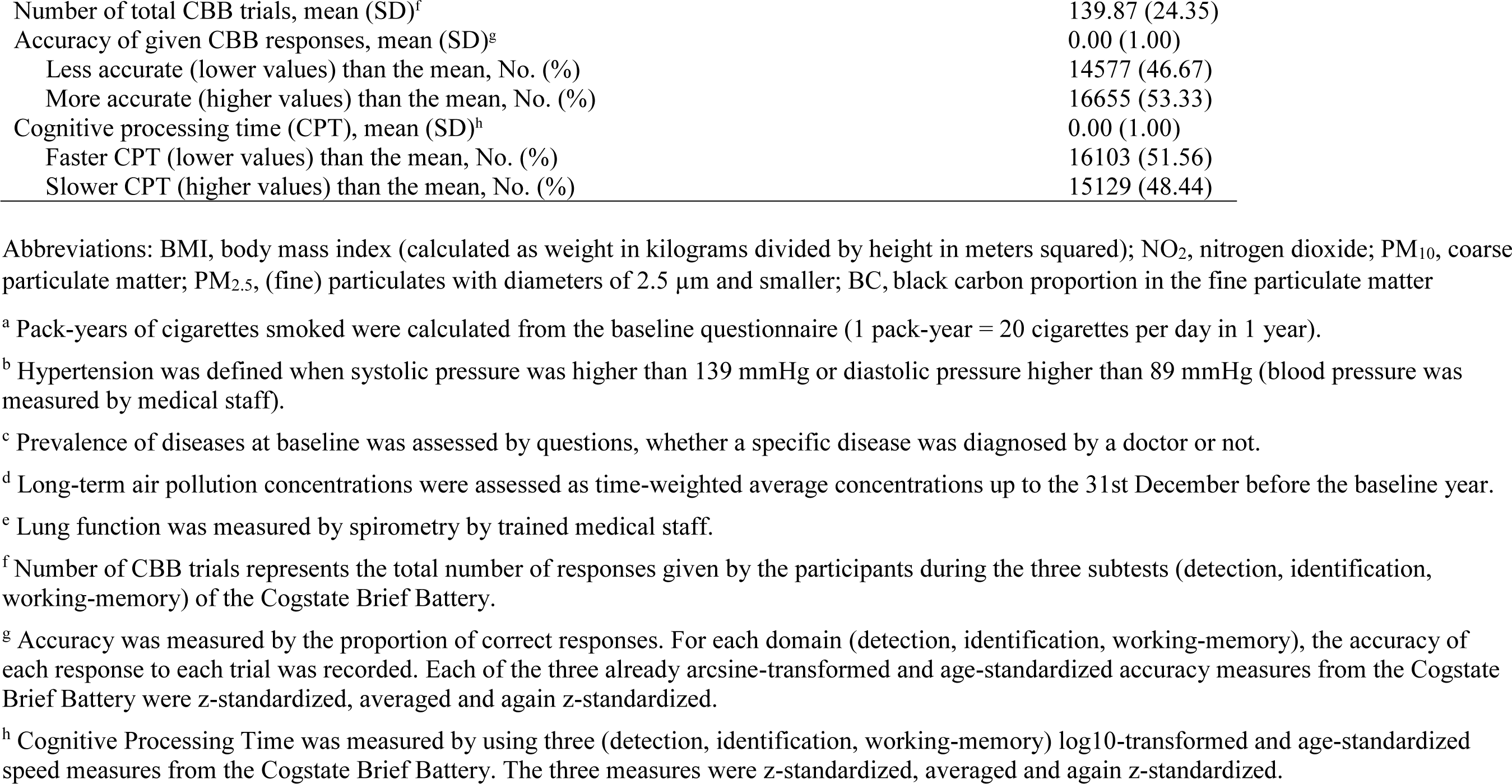
Descriptive Statistics of the Study Participants (n = 31232) up to Baseline, at Baseline (2006-2012) and at Follow-Up (2014-2015) in the Study Population Based on the Dutch Lifelines Cohort Study

**Figure 2.**
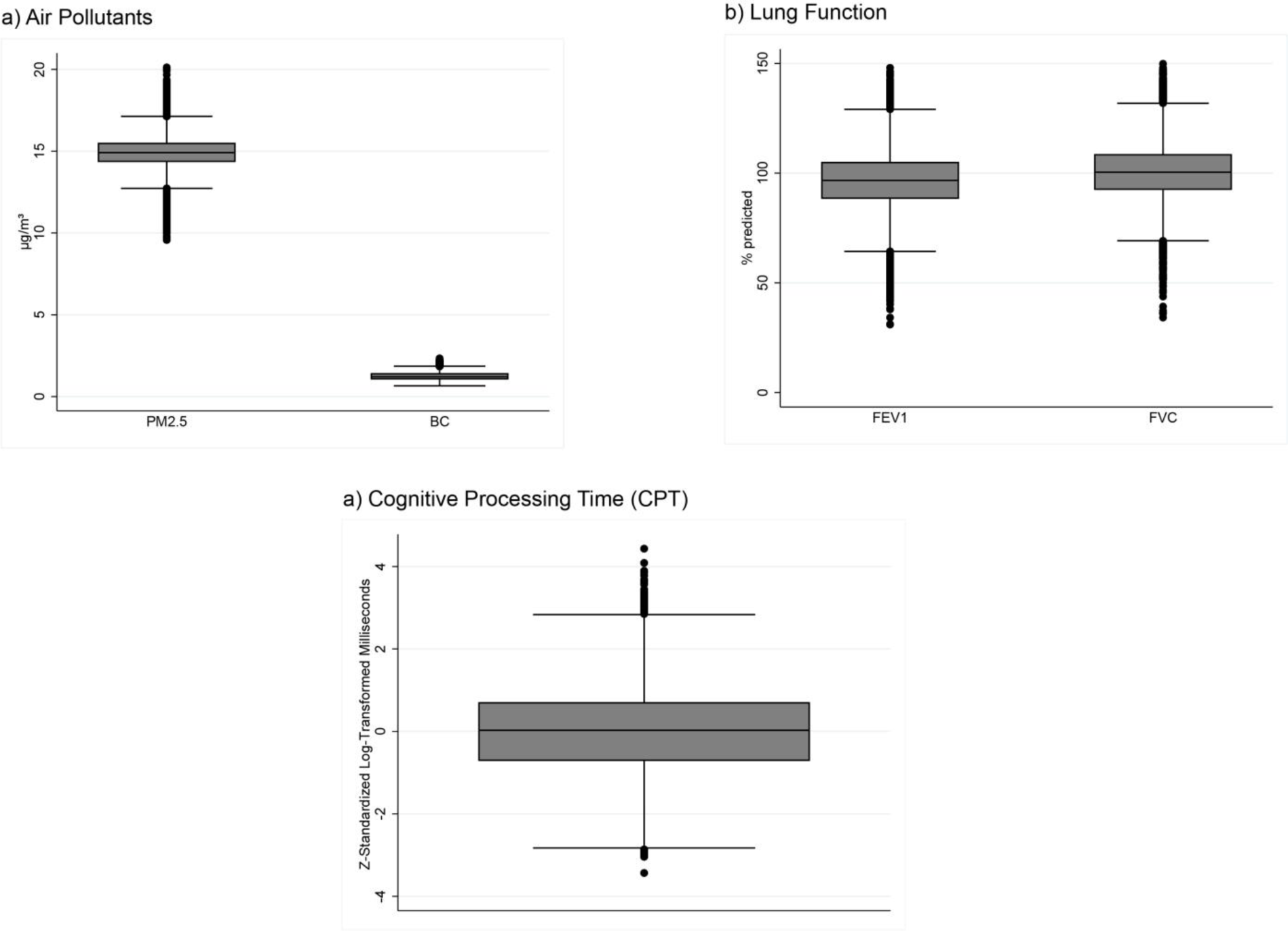
Distribution of Long-Term Exposure to PM_2.5_ and Black Carbon up to Baseline, Lung Function at Baseline (2006-2012) and Cognitive Processing Time at Follow-Up (2014-2015) Boxplots included average time-weighted long-term air pollution concentrations a) of the study participants up to baseline, b) lung function measures at baseline (2006 to 2012), and c) cognitive processing time (CPT), the outcome of interest, at follow-up (2014 to 2015). The boxes indicate the interquartile range (IQR) and the line in the center indicates the median concentration. Whiskers extend to 1.5 times the IQR to the most distant observation within that distance. Outlier observations are shown as circles. Exposure data a) are shown by air pollutants, namely fine particulate matter (PM_2.5_) and black carbon proportion (BC). Lung function data b) are shown by spirometry measure, namely the forced expiratory volume in 1 second (FEV_1_) and the forced volume capacity (FVC). c) shows the data of the cognitive processing time (CPT). Higher values indicate slower CPT and worse cognitive performance, and lower values faster CPT and better cognitive performance.

There were 14895 (47.69%) people who had zero pack-years of cigarettes smoked, suggesting that they were never-smokers, and the smokers had an average of 10.93 pack-years. The body mass index of 112254 (39.24%) participants indicated they were overweight (BMI ≥ 25), and 3952 (12.65%) were obese (BMI ≥ 30). 7.43% of the participants had prevalent asthma at baseline diagnosed by a doctor, 4.30% COPD, 6.58% diabetes, 10.11% depression, and 0.45% had ever a stroke before. Age at baseline ranged from 18 to 85 and the average age was 43.95 years.

### Associations between Air Pollution Exposure and Cognitive Processing Time (CPT)

First, we estimated the total effect showing the overall associations between long-term air pollution exposure up to baseline and CPT at follow-up (2014-2015). We found that higher exposure to PM_2.5_ was significantly related to slower CPT and so worse cognitive performance [18.33×10^−3^ SD above the mean (95% CI: 6.84, 29.81)] (**Figure 3, Supplementary Table S1**). We additionally estimated adjusted CPT predictions for the values of PM_2.5_ from the minimum integer value (9) to the maximum (20) value under our model assumptions (**Supplementary Figure S3**). This showed that up to an average exposure level of 15µg/m^3^ [0,87×10^−3^ SD above the mean (95% CI: −9.78, 29.81)] no effect of PM_2.5_, on CPT was seen. From an average value of 16µ/m^3^ [18.67×10^−3^ SD above the mean (95% CI: 2.82, 24.92)] onwards, however, PM_2.5_ had an adverse effect on cognition and was associated to slower (values > 0) z-standardized CPT. For BC, however, no significant associations existed.

**Figure 3.**
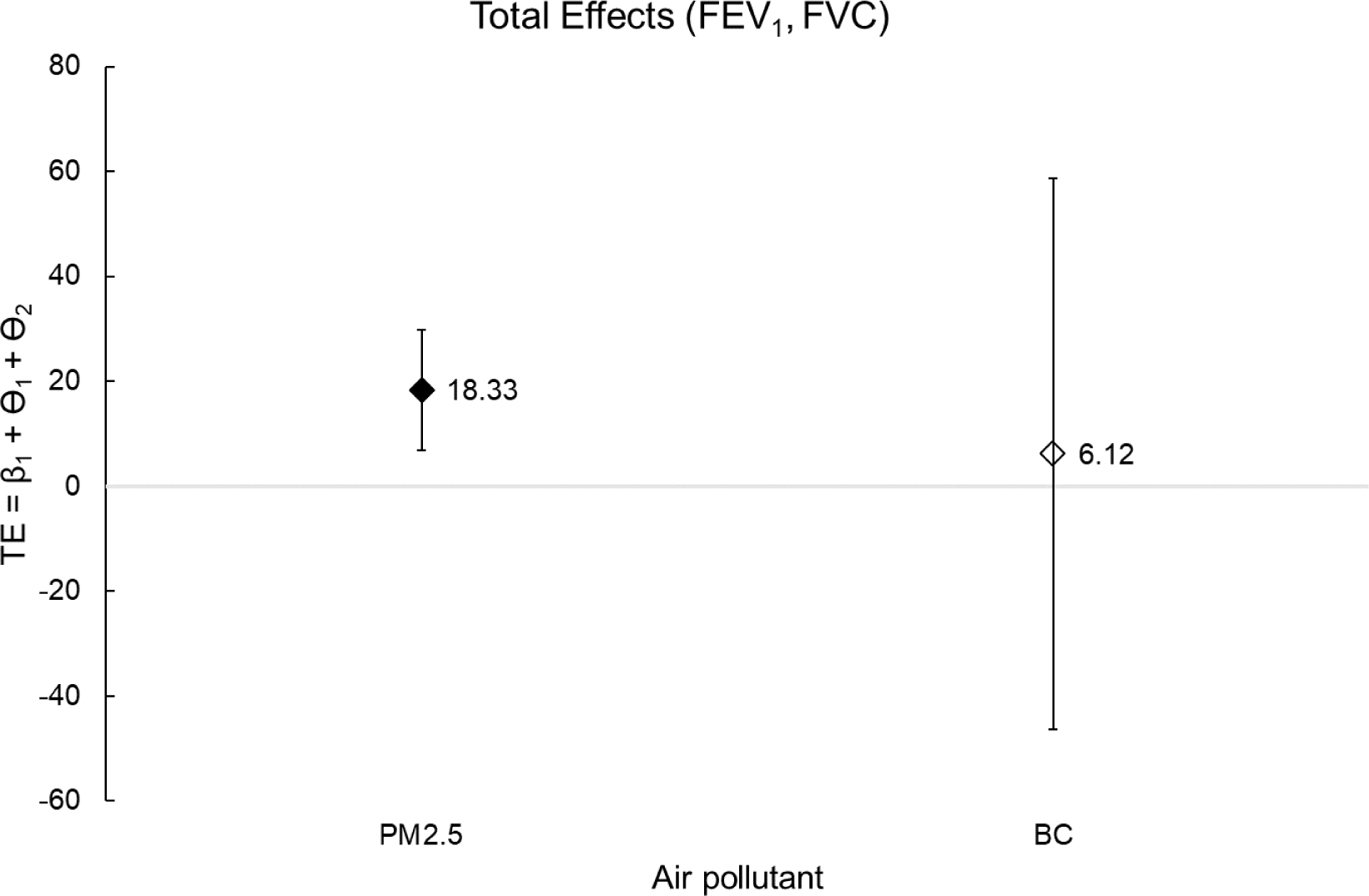
Total Effects **-** Associations between Long-Term Exposure to PM_2.5_ and BC, and Cognitive Processing Time (CPT) Abbreviations: PM_2.5_, (fine) particulates with diameters of 2.5 µm and smaller; BC, black carbon proportion in the fine particulate matter; FEV_1_, Forced Expiratory Volume in one second; FVC, Forced Volume Capacity Interval plots show the Total Effects (a, b) of the associations between long-term time-weighted air pollution (in µg/m^3^) up to baseline and Cognitive Processing Time (CPT) measured by the Cogstate Brief Battery. Linear structural equation models with robust standard errors by Huber/ White were performed. Illustrated were the point estimators and confidence intervals coming from single models for each air pollutant, controlled for sex, age, province of residence, educational level, income, pack-years of cigarettes smoked, hypertension, BMI, the age- and z-standardized CBB accuracy, and the total number of CBB trials. Shown values for point estimators, lower and upper confidence intervals were multiplied by 10^3^.

### Mediation Analysis and Decomposition of the Total Effect into the Direct and Indirect Effect

Second, we conducted a decomposition of the total effects into a direct and an indirect effect. The direct effect represents the direct route of air pollutants through the olfactory nerve or the lung, with subsequent entry into the blood stream, providing access to the brain. The direct effects for PM_2.5_ on CPT were seen even when we controlled for both lung function measures at baseline (2006-2012), so the indirect routes, namely for FEV_1_ [18.00×10^−3^ SD above the mean (95% CI: 6.51, 29.49)] and FVC [17.85×10^−3^ SD above the mean (95% CI: 6.35, 29.34)]. No significant direct effects were found for BC (**Figure 4**).

**Figure 4.**
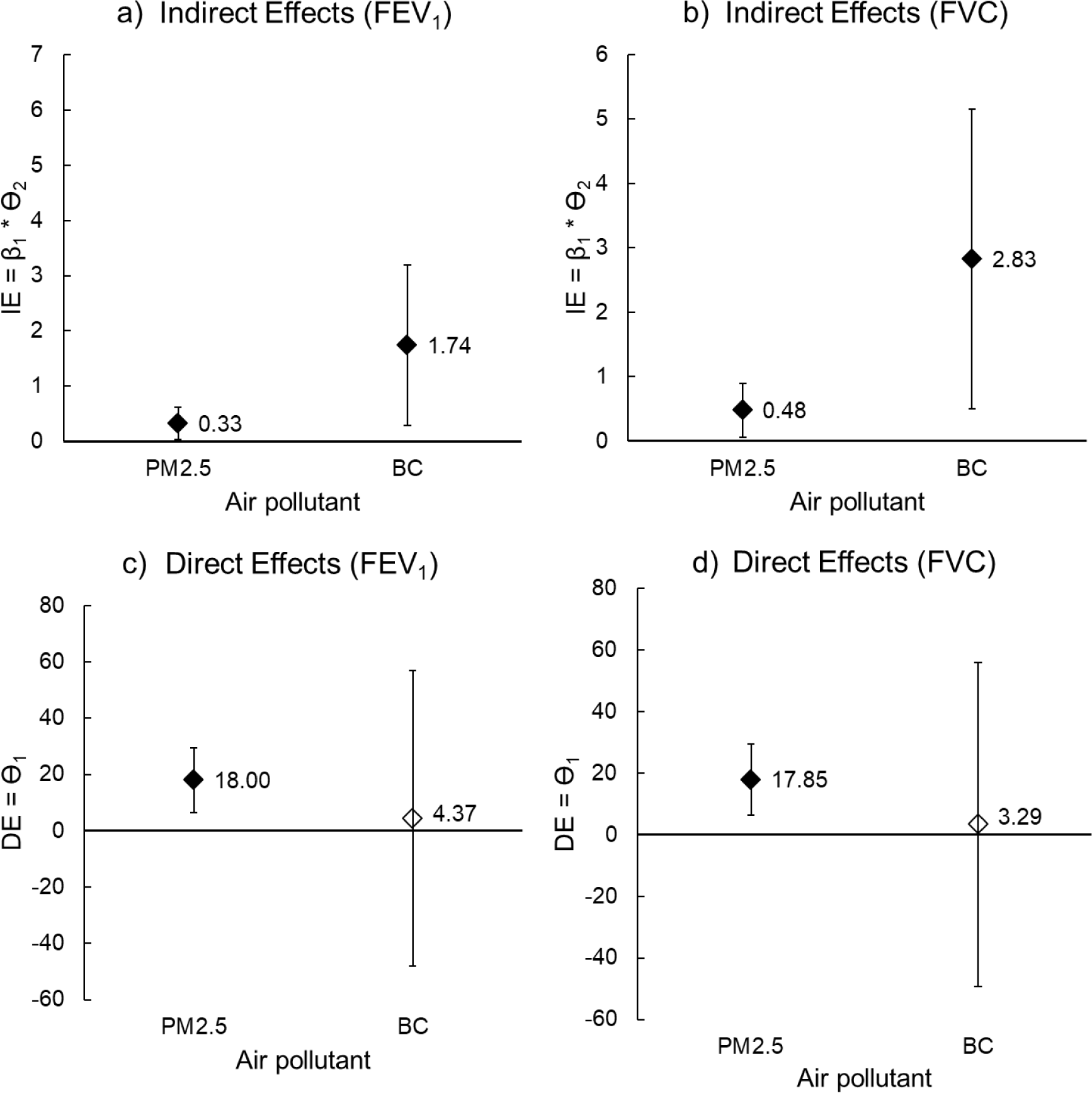
Decomposition of the Associations between Long-Term Exposure to PM_2.5_ and Black Carbon and Cognitive Processing Time (CPT) Abbreviations: PM_2.5_, (fine) particulates with diameters of 2.5 µm and smaller; BC, black carbon proportion in the fine particulate matter; FEV_1_, Forced Expiratory Volume in one second; FVC, Forced Volume Capacity Interval plots show the Total Effects (A, B) and their decompositions of the associations between long-term time-weighted air pollution (in µg/m^3^) up to baseline and Cognitive Processing Time (CPT) measured by the Cogstate Brief Battery. The direct c), d) and the indirect a), b) routes over the lung function measures FEV_1_ and FVC (potential mediators) at baseline (2006-2012) were modelled simultaneously by performing linear structural equation models with robust standard errors by Huber/ White. Illustrated were the point estimators and confidence intervals coming from single models for each air pollutant, controlled for sex, age, province of residence, educational level, income, pack-years of cigarettes smoked, hypertension, asthma, COPD, diabetes, depression, stroke, multiple sclerosis, BMI, the age- and z-standardized CBB accuracy, and the total number of CBB trials. Shown values for point estimators, lower and upper confidence intervals were multiplied by 10^3^.

The indirect effect represents the indirect route of air pollutants, which first impair the lung, which in turn leads to worse cognitive performance. For PM_2.5_, we observed mediation by both lung function measures, namely for FEV_1_ [0.33×10^−3^ SD above the mean (95% CI: 0.04-0.22)] and FVC [0.44×10^−3^ SD above the mean (95% CI: 0.06-0.90)] (**Figure 4** and see **Supplementary Table S1** for the estimation parameters of *β*_1_, *θ*_1_ and). We also found indirect effects for BC, namely for both lung function measures (FEV_1_: [1.74×10^−3^ SD above the mean (95% CI: 0.29, 3.20); FVC: [2.83 (95% CI: 0.51, 5.15)].

Our first sensitivity analysis using g-formula supported our results and showed that our model approach came to nearly the same estimations (**Supplementary Table S2**). The second sensitivity analysis including participants aged 45 and older only (see **Supplementary Table S4** for descriptive statistics) showed also same results than the all-participant model in line with our hypothesis (**Table 2**). However, we did not find an indirect effect of PM_2.5_ by FEV_1_ due to a lack of significance. In the third sensitivity analysis, we used the average concentrations of PM_2.5_ and BC at residential address of participants at baseline only (see **Supplementary Table S4** for descriptive statistics), and we found same results as in our model with time-weighted average concentrations over at least ten years before baseline (**Table 2**). This showed that our sample selection procedure and the inclusion of participants with at least ten years of residential address history available did not bias the results.

**Table 2.**
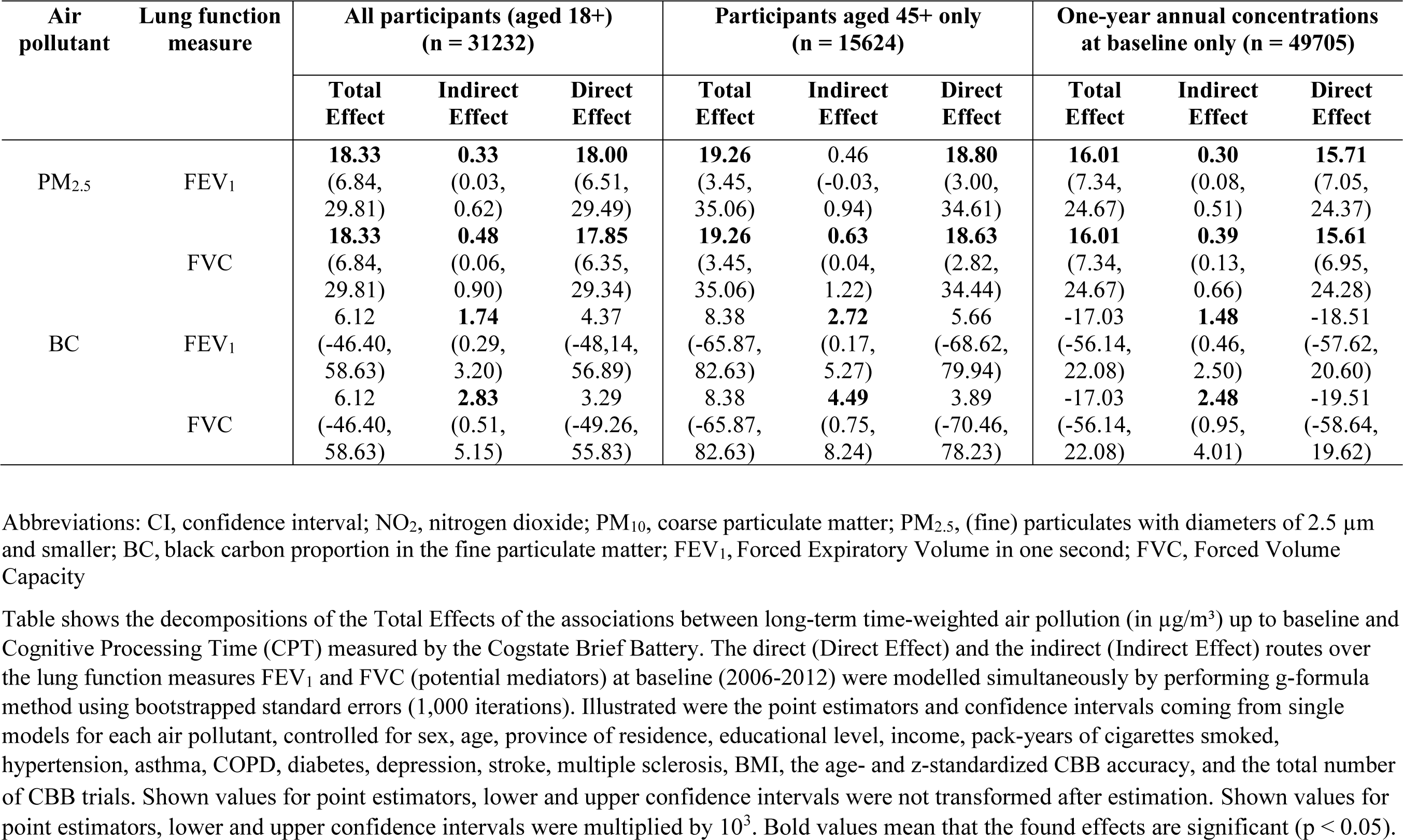
Outcomes of our decomposition analysis for our main analysis, and two sensitivity analyses: one using participants aged 45+ only, and one using one-year annual concentrations of the air pollutant at baseline only, compared to using long-term exposure

| Air pollutant | Lung function measure | All participants (aged 18+) (n = 31232) |  |  | Participants aged 45+ only (n = 15624) |  |  | One-year annual concentrations at baseline only (n = 49705) |  |  |
| --- | --- | --- | --- | --- | --- | --- | --- | --- | --- | --- |
|  |  | Total Effect | Indirect Effect | Direct Effect | Total Effect | Indirect Effect | Direct Effect | Total Effect | Indirect Effect | Direct Effect |
| PM<sub>2.5</sub> | FEV<sub>1</sub> | **18.33**<br>(6.84, 29.81) | **0.33**<br>(0.03, 0.62) | **18.00**<br>(6.51, 29.49) | **19.26**<br>(3.45, 35.06) | 0.46<br>(-0.03, 0.94) | **18.80**<br>(3.00, 34.61) | **16.01**<br>(7.34, 24.67) | **0.30**<br>(0.08, 0.51) | **15.71**<br>(7.05, 24.37) |
|  | FVC | **18.33**<br>(6.84, 29.81) | **0.48**<br>(0.06, 0.90) | **17.85**<br>(6.35, 29.34) | **19.26**<br>(3.45, 35.06) | **0.63**<br>(0.04, 1.22) | **18.63**<br>(2.82, 34.44) | **16.01**<br>(7.34, 24.67) | **0.39**<br>(0.13, 0.66) | **15.61**<br>(6.95, 24.28) |
| BC | FEV<sub>1</sub> | 6.12<br>(-46.40, 58.63) | **1.74**<br>(0.29, 3.20) | 4.37<br>(-48.14, 56.89) | 8.38<br>(-65.87, 82.63) | **2.72**<br>(0.17, 5.27) | 5.66<br>(-68.62, 79.94) | -17.03<br>(-56.14, 22.08) | **1.48**<br>(0.46, 2.50) | -18.51<br>(-57.62, 20.60) |
|  | FVC | 6.12<br>(-46.40, 58.63) | **2.83**<br>(0.51, 5.15) | 3.29<br>(-49.26, 55.83) | 8.38<br>(-65.87, 82.63) | **4.49**<br>(0.75, 8.24) | 3.89<br>(-70.46, 78.23) | -17.03<br>(-56.14, 22.08) | **2.48**<br>(0.95, 4.01) | -19.51<br>(-58.64, 19.62) |

Abbreviations: CI, confidence interval; NO<sub>2</sub>, nitrogen dioxide; PM<sub>10</sub>, coarse particulate matter; PM<sub>2.5</sub>, (fine) particulates with diameters of 2.5 µm and smaller; BC, black carbon proportion in the fine particulate matter; FEV<sub>1</sub>, Forced Expiratory Volume in one second; FVC, Forced Volume Capacity
Table shows the decompositions of the Total Effects of the associations between long-term time-weighted air pollution (in µg/m<sup>3</sup>) up to baseline and Cognitive Processing Time (CPT) measured by the Cogstate Brief Battery. The direct (Direct Effect) and the indirect (Indirect Effect) routes over the lung function measures FEV<sub>1</sub> and FVC (potential mediators) at baseline (2006-2012) were modelled simultaneously by performing g-formula method using bootstrapped standard errors (1,000 iterations). Illustrated were the point estimators and confidence intervals coming from single models for each air pollutant, controlled for sex, age, province of residence, educational level, income, pack-years of cigarettes smoked, hypertension, asthma, COPD, diabetes, depression, stroke, multiple sclerosis, BMI, the age- and z-standardized CBB accuracy, and the total number
of CBB trials. Shown values for point estimators, lower and upper confidence intervals were not transformed after estimation. Shown values for point estimators, lower and upper confidence intervals were multiplied by $10^3$ . Bold values mean that the found effects are significant ( $p < 0.05$ ).

By calculating the importance of direct and indirect effects, we found that about 98% (FEV_1_) or 97% (FVC) of the total effect of PM_2.5_ was directly associated with cognitive performance. The highest indirect effect proportions were seen for BC (FVC, 46.22%) and the lowest indirect effect proportion was seen for PM_2.5_ by FEV_1_ (1.78%) (**Figure 5**).

**Figure 5.**
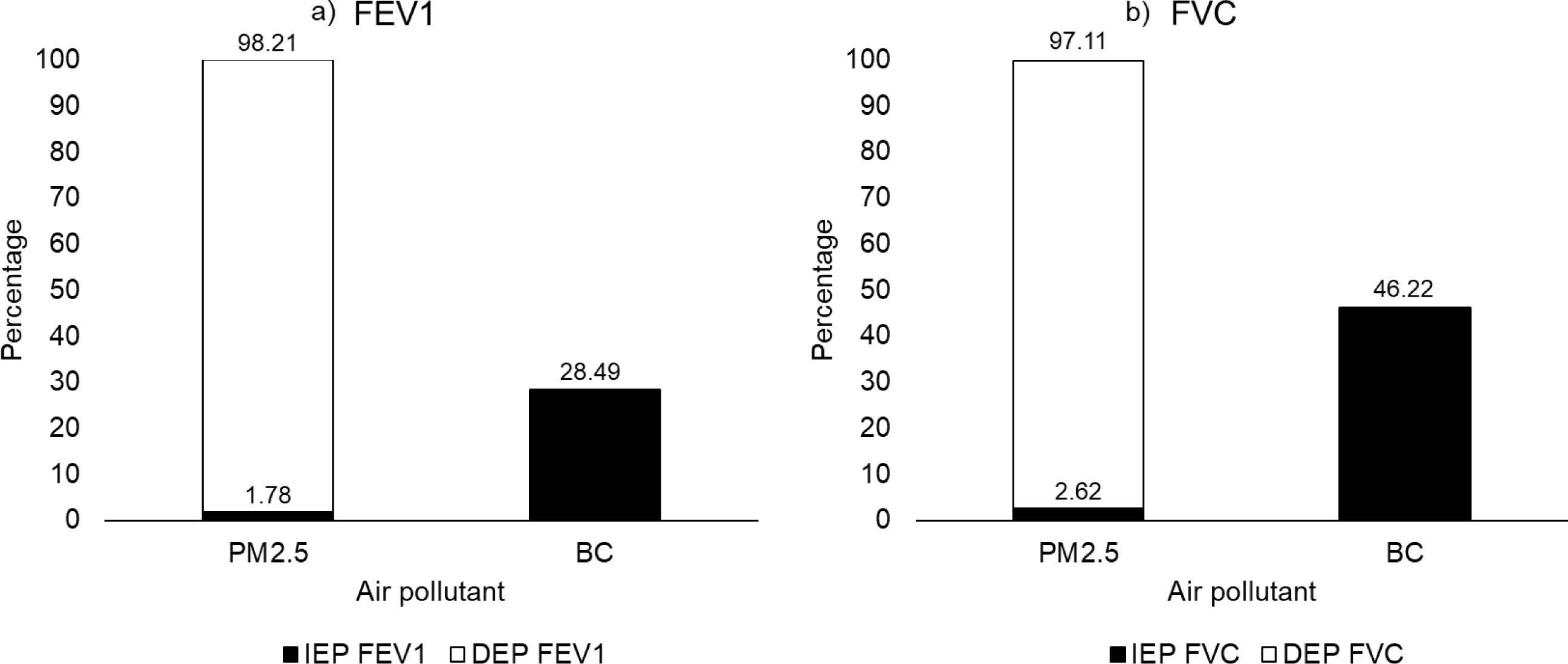
The Importance of the Found Indirect (IEP) and Direct Effects (DEP) of Long-Term Exposure to PM_2.5_ and BC on Cognitive Processing Time (CPT) Calculated as Proportions in Percentage of the Total Effects Abbreviations: PM_2.5_, (fine) particulates with diameters of 2.5 µm and smaller; BC, black carbon proportion in the fine particulate matter; FEV_1_, Forced Expiratory Volume in one second; FVC, Forced Volume Capacity; IEP, indirect effect proportion; DEP, direct effect proportion Bars show the found significant indirect effects (IEP) and the direct effect proportions (DEP) found in the decomposition of the total effects for both lung function measures FEV_1_ a) and FVC b) acting as potential mediators. Indirect effect proportions were calculated by dividing the size of the indirect effects by their total effects. The direct effect proportions were calculated by dividing the size of the direct effects by their total effects. The models were controlled for sex, age, province of residence, educational level, income, pack-years of cigarettes smoked, hypertension, asthma, COPD, diabetes, depression, stroke, multiple sclerosis, BMI, the age- and z-standardized CBB accuracy, and the total number of CBB trials.

### The influence of Confounders on the Associations between Air Pollution and Cognitive Performance

Turning to the covariates included in our study, we found that age was not correlated with cognitive function [−0.05×10^−3^ SD below mean (95% CI: −1.17, 1.07)] indicating successful age-standardization of our outcome of interest, CPT. As expected, higher educated people had better cognitive performance [−503.26×10^−3^ SD below mean (95% CI: −733.03, −273.50)] than participants without any educational level. We further found that prevalent diabetes was related to poor cognition [−99.12×10^−3^ SD below mean (95% CI: −54.12, −144.12)]. The other morbidities showed no significant effect. Furthermore, women fared worse than men [−68.78×10^−3^ SD below mean (95% CI: −91.34, −46.22)].

We found, as expected, that participants with prevalent COPD [FEV_1_: −6.87% predicted (95% CI: −7.71, −6.04); FVC: −3.36% predicted (95% CI: −4.06, −2.66)] and asthma [FEV_1_: −4.54% predicted (95% CI: −5.11, −3.97); FVC: −1.13% predicted (95% CI: −1.64, − 0.63)] had worse lung function. But also diseases, which were not directly related to respiratory health, were associated with worse lung function, namely stroke [FEV_1_: −4.43% predicted (95% CI: −6.63, −2.22); FVC: −3.36% predicted (95% CI: −5.25, −1.02)], diabetes [FEV_1_: −0.70% predicted (95% CI: −1.28, −0.13); FVC: −1.21% predicted (95% CI: −1.76, − 0.66)], and hypertension [FEV_1_: −1.24% predicted (95% CI: −1.66, −0.83); FVC: −1.19% predicted (95% CI: −1.57, −0.80)].

We additionally found that lower income, a higher number of packyears and obesity, but not a lower educational level, were associated with lower FEV_1_ and FVC. We observed that higher age was related to both higher FEV_1_ and FVC indicating that the age-standardization by the GLI reference values was not fully successful and an additional inclusion of age in our models was important to avoid confounding by age. For sex, however, we found no association with lung function confirming that the sex-standardization was successful.

## DISCUSSION

### Summary of the Findings

To the best of our knowledge, our cohort study is one of the first to demonstrate the importance of the direct and the indirect route over the lung taken by inhaled fine particulate matter in general and Black Carbon in particular in cognitive impairments for both genders.

In the low pollution setting of the Netherlands, higher exposure to PM_2.5_ was associated with slower cognitive processing time at follow-up (2014-2015) among participants aged 18+, and associations were also evident in a sensitivity model including participants aged 45 and older. Further analyses showed that an average PM_2.5_ exposure level of 16 to 20 µg/m^3^ was significantly related to worse age-standardized cognitive performance, whereas people experiencing only 9 to 14 µg/m^3^ on average had better cognitive performance. By decomposing the total effects into a direct and an indirect effect respectively, we showed that PM_2.5_ was directly related to slower cognitive processing time and, thus to worse cognitive performance. This direct effect constituted more than 97% of the total effect of PM_2.5_ on cognitive performance when we included mediation by lung function at baseline (2006-2012) in our model. In addition to the direct effect, there was also small significant indirect effects for both lung function measures FEV_1_ and FVC contributing just about 2 to 3% to the total effect.

For BC, the associations were solely mediated via lung function. We found significant indirect effects, which contributed about 28% (FEV_1_) to 46% (FVC) to the total effects. Both mediations however, were too small to translate significance to the total effect since there were insignificant direct effects which contribute (partially) to the total effect as well.

As most important covariates/ confounders, we identified educational level and diabetes prevalence for cognitive performance, and prevalence of COPD, asthma, stroke, diabetes, and hypertension as well as income and packyears for lung function.

The main aim of this study was to test the two hypothesized routes taken by fine particulate matter in general (PM_2.5_) and Black Carbon (BC) as one of the main components of the fine particulate matter. Our results indicate that the direct route may be more important for PM_2.5_ and the indirect route for BC.

### Interpretation and Comparison of the Findings

Turning to the interpretation of our findings, our results suggest that BC may not be able to cross the direct route to reach the brain among participants aged 18 and older. Previous research has, in contrary to our results, found that higher exposure to BC was related to worse cognitive functioning (Colcino et al. 2017; Wurth et al. 2018). These studies, however, included people aged 60 and older only. An explanation could be that older people (≥ 65 years old), who are more vulnerable and fragile, are more prone to the negative effects of air pollution (Shumake et al. 2013; Simoni et al. 2015) than younger people.

We found that PM_2.5_, however, may affect cognition directly. It is known that fine particles, e.g. PM_2.5_ and especially ultrafine particles (diameter < 0.01µm), a component of PM_2.5_ as well, are potentially able to penetrate the central nervous system, resulting in subsequent neuroinflammation (Maher et al. 2016). We further know that microglia may be activated by the direct route, but also upon occurrence of systemic neuroinflammation (Widmann and Heneka 2014). In vivo and in vitro studies also provide insights into the neurotoxic effect of PM exposure; i.e., higher levels of PM are linked to significantly higher levels of pro-inflammatory cytokines such as interleukin, as well as tumor necrosis factor and glial responses indicating the presence of inflammation (Kilian and Kitazawa 2018). In turn, neuroinflammation is an important risk factor for neurodegenerative diseases, e.g. Alzheimer’s and Parkinson’s disease (Glass et al. 2010; Heneka et al. 2015; Sarlus and Heneka 2017). Another possibility is that the pollutants inhaled reach the brain via the lung, which provides the initial entry to the body, and then pass from the alveoli into the bloodstream, using circulation to reach the central nervous system (CNS) (Lee and Shah 2018). We further know that exposure to PM_2.5_ is associated with DNA methylation, which may in turn affect lung function or/ and brain health negatively (Shi et al. 2019).

Additionally, we found evidence for the relevance of the indirect route over the lung in cognitive impairments by fine particulate matter. That is, PM_2.5_ to a small extent and BC to a greater extent affect cognitive performance by first impairing the lung function and subsequently leading to cognitive impairment. For BC, this seemed to be the main route in our study to unfold adverse cognition effects, since we found that the effect of BC on cognitive performance was solely mediated by FEV_1_ and FVC with a mediated proportion of 28 to 46% and we found no evidence that BC is able follow the direct route. Previous studies have also found that higher exposure to BC and PM_2.5_ are associated with lower FEV_1_ and FVC (Guo et al. 2018; Lepeule et al. 2014), and inflammatory processes in the lung, oxidative stress and activation of microglia cells resulting in subsequent neuroinflammation have been suggested as causal pathways (Guarnieri and Balmes 2014). However, the fact that the found mediated proportion is only 28 to 46% for BC suggests that there are potential additional routes missed in our study, e.g. cardiovascular health/ diseases, which could be relevant in impairing cognition by BC. We know accordingly from previous research that higher exposure to BC was related to adverse cardiovascular outcomes (Gan et al. 2011). In turn, another study has found that better cardiovascular health was related to better cognitive health (Kulshreshtha et al. 2019).

Our study was conducted in the low-pollution setting of adults aged 18+ in the Netherlands. For PM_2.5_ (14.95 µg/m^3^), the exposure level was clearly lower compared to the relatively restrictive EU-wide limit values (25 µg/m^3^ for PM_2.5_) (Guerreiro et al. 2018), and also lower compared to previous epidemiological cohort studies (ECRHS, 16 µg/m^3^; EGEA, 15 µg/m^3^; E3N, 15 µg/m^3^; SALIA, 18 µg/m^3^; SAPALDIA, 17 µg/m^3^) (Jacquemin et al. 2015). This was also true for BC, which was 1.25 µg/m^-1^ in our study, but 1.5 µg/m^-1^ or higher in other cohorts (ECRHS, 2.0 µg/m^-1^; EGEA, 2.1 µg/m^-1^; E3N, 1.8 µg/m^-1^; SALIA, 1.5 µg/m^-1^; SAPALDIA, 1.9 µg/m^-1^) (Jacquemin et al. 2015). Higher exposure levels in other spatial units or countries may result in even stronger associations between air pollution and cognition than what is demonstrated in this study’s population, under the assumption of a non-linear, e.g exponential, dose-response relationship. To the best of our knowledge, there is no previous study which evaluated the dose-response relation between fine particulate matter and cognitive performance among adults, so that more research is needed here.

### Strengths and Limitations

Our study has several strengths. First, we did not only explore the overall relationship (total effect) between fine particulate matter and cognitive performance as done by previous research (Power et al. 2011; Wurth et al. 2018; Zhang et al. 2018), but additionally performed a decomposition of the total effects in order to disentangle the potential routes taken by the air pollutants, which are closely related to causal mechanisms/ pathways. For this purpose, we applied a causal mediation approach by using SEM. To the best of our knowledge, the only previous cohort study that explored the mediating role of lung function was not able to find any significant indirect effects (Hüls et al. 2018), potentially because they observed only a small female cohort in Germany. The adverse total effect of PM_2.5_ they observed, however, supports our results.

Second, we calculated time-weighted average concentrations over a minimum of ten years up to baseline to measure the long-term exposure to air pollutants. Using one-year concentrations only, as done in previous studies (Colcino et al. 2017; Nußbaum et al. 2020; Wang et al. 2020; Wurth et al. 2018), may confound the results by unobserved positive health selection into living environments with low exposure levels (Oakes 2014), even when our third sensitivity model showed that this is not true in our study.

Third, we used a standardized and multidimensional (composite) outcome variable, cognitive processing time (CPT), which was computed in close contact with the Cogstate research team, to measure the participants’ cognitive performance. The speed measures coming from the CBB may be summarized into a composite, also used to determine the overall cognitive performance and not only to detect clinical abnormalities. Previous studies mostly used the MMSE as an outcome to measure mild cognitive impairment, e.g. when MMSE scores were ≤ 25 (Colcino et al. 2017; Shehab and Pope 2019; Wang et al. 2020). This, however, has a relative lack of sensitivity when detecting subtle cognitive impairments (Proust-Lima et al. 2007) and detailed differences in cognitive performance within a specific population.

Fourth, a series of sensitivity analyses confirmed our results. (1) A replication analyses using g-formula and bootstrapping method for testing the causal routes (**Supplementary Table S2**), yielded similar results to our SEM approach. (2) This was also true for the analysis of individuals aged 45 and older only, which gives even more plausibility to our results, because previous research suggested that cognitive decline starts from the mid-40s (Singh-Manoux et al. 2012). (3) Using one-year air pollution concentrations at baseline only, did not change our results underlining that our sample selection procedure of including individuals with a residential history of ten years only did not bias our findings.

Despite the important strengths of our study, there are some limitations. First, ambient air pollution concentrations at the residential address, especially those estimated by land use regression models, do not reflect the overall air pollution people are indeed exposed to in their living environments. People are also exposed to (potentially different) air pollution levels, which are relevant for brain health, during daily road travel or when they are indoor (Saenz et al. 2018). Second, we used the air pollution exposure data from the ELAPSE project, which seems to have better quality than previous data (de Hoogh 2018). But, the data were only available for the year 2010 and these data from 2010 were linked to the participants address history, so that the calculated average long-term exposure concentrations at home used in this study just act as a proxy for the real air pollution dose participants experienced in their surroundings. We feel however, that our way to handle the data should deliver at least better results than using annual concentration from one year at baseline only. Third, we analyzed only the levels of health and not the changes in health over time, which suggests generally more causal explanatory power. We, however, minimized the effect of this limitation by ensuring correct causal time order between cause, mediator and outcome. Furthermore, we included only those participants for whom exposure data was available for at least ten years, and verified our results by performing causal mediation analyses with a potential outcome approach (see our first sensitivity analysis). Fourth, we were not able to account for genetic factors, e.g. the apolipoprotein-ε4 allele (APOE-ε4), which are known to be the most prevalent genetic risk factors for Alzheimer’s disease (Miyata and Smith 1996), and whose variants modify the association between air pollution and cognitive impairment resulting in a stronger air pollution effect for APOE risk variant carriers (Schikowski et al. 2015). Fifth, our models assumed a linear dose/-response relationship between air pollution exposure and cognitive performance. In doing so, we followed previous studies in the field, which explored e.g. the dose-response relationship between PM_2.5_ and daily deaths (Schwartz et al. 2002), PM_2.5_ and daily respiratory deaths (Ren et al. 2017), prenatal exposure to PM_2.5_ and development of brain white matter and cognition in later childhood (Bradley et al. 2015), which all identified a linear relationship. However, there is also evidence for the existence of non-linear relations between PM_2.5_ and mortality due to respiratory disease (COPD, lung cancer, lower respiratory infection), especially for lower concentrations below 25 µg/m^3^ (Cohen et al. 2017).

## Conclusion

Our study provides new insights in the association between ambient exposure to fine particulate matter and brain health by disentangling underlying direct and indirect pathways over lung function. Our results emphasize the importance of the lung acting as a mediator in this relationship. Especially BC seems to impair the lung or activate inflammatory responses of immune cells residing in the lung, causing subsequent cognitive impairments. Thus, brain health seems to depend on, among others, a good lung function, and lung function in turn may benefit from low air pollution exposure. Fine particles as PM_2.5_ seem to mainly follow the direct route, which calls for a special attention to fine particles due to their ability to reach vital organs directly. Future research is needed that explores, beside of the indirect lung function route found in our study, further pathways taken by inhaled air pollutions to subsequently affect cognition.

## Data Availability

The data analyzed in this study was obtained from the Lifelines biobank (project application number OV15_0307). Requests to access this dataset should be directed to Lifelines Research Office.

## Acknowledgements

We thank Lifelines for providing the data and their support. The Lifelines Biobank initiative was made possible by a subsidy from the Dutch Ministry of Health, Welfare and Sport, the Dutch Ministry of Economic Affairs, the University Medical Center Groningen (UMCG the Netherlands), University Groningen and the Northern Provinces of the Netherlands. We further thank the Cogstate Research Team for their support in handling the data coming from the Cogstate Brief Battery, and Renée Lüskow for her language editing.

We have no conflict of interest to declare.

## Figure Legends

**Figure 1**. Routes Taken by Inhaled Fine Particles to Cause Subsequent Cognitive Impairments

**Figure 2**. Distribution of Long-Term Exposure to PM_2.5_ and Black Carbon up to Baseline, Lung Function at Baseline (2006-2012) and Cognitive Processing Time at Follow-Up (2014-2015)

**Figure 3**. Total Effects **-** Associations between Long-Term Exposure to PM_2.5_ and BC, and Cognitive Processing Time (CPT)

**Figure 4**. Decomposition of the Associations between Long-Term Exposure to PM_2.5_ and Black Carbon and Cognitive Processing Time (CPT)

**Figure 5**. The Importance of the Found Indirect (IEP) and Direct Effects (DEP) of Long-Term Exposure to PM_2.5_ and BC on Cognitive Processing Time (CPT) Calculated as Proportions in Percentage of the Total Effects

