## Supplemental Material for "Long-Term Exposure to Fine Particulate Matter, Lung Function and Cognitive Performance: A Prospective Dutch Cohort Study on the Underlying Routes"

**Supplementary Table S1.** Decomposition of the Associations between Time-Weighted Average, Long-Term Exposure to $PM_{2.5}$ and Black Carbon and Cognitive Processing Time (CPT) Showing Single Parameters among the Entire Study Population

**Supplementary Table S2.** Sensitivity Analysis of the Model Approach – Causal Mediation Analyses Using G-Formula and Bootstrapping Method

**Supplementary Table S3.** Comparison of the Descriptive Statistics among Participants with Complete (n = 31232) and Incomplete Data (n = 110908)

**Supplementary Table S4.** Descriptive Statistics - Sensitivity Models among Participants Aged 45+ Only and by Using One-Year Annual Concentrations Only

**Supplementary Figure S1.** Construction of the Study Population Based on the Lifelines Cohort Study Population Aged 18+

**Supplementary Figure S2.** Used Structural Equation Model (SEM) to Test the Direct and Indirect Routes of Air Pollutants on Cognitive Processing Time (CPT)

**Supplementary Figure S3.** Adjusted Predictions of Cognitive Processing Time for the Total Effects of $PM_{2.5}$

**Supplementary Figure S4.** Distribution of the Mediator Variable ($FEV_1$)

**Supplementary Figure S5.** Distribution of the Mediator Variable (FVC)

**Supplementary Figure S6.** Distribution of the Outcome Variable (CPT)

This supplementary material has been provided by the authors to give readers additional information about their work.

**Supplementary Table S1.** Decomposition of the Associations between Time-Weighted Average, Long-Term Exposure to $PM_{2.5}$ and Black Carbon and Cognitive Processing Time (CPT) Showing Single Parameters among the Entire Study Population

| **Air pollutant** | **Lung function measure** | **Estimations** | | |
|---|---|---|---|---|
| | | $\beta_1$ | $\theta_2$ | $\theta_1$ |
| $PM_{2.5}$ | $FEV_1$ | **-0.2485201**<br>(-0.3960919, -0.1009484) | **-0.0013092**<br>(-0.0021924, -0.0004259) | **0.0180010**<br>(0.0065133, 0.0294888) |
| | FVC | **-0.4211821**<br>(-0.5628614, -0.2795029) | **-0.0011381**<br>(-0.0020566, -0.0002196) | **0.0178471**<br>(0.0063529, 0.0293412) |
| BC | $FEV_1$ | **-1.3064010**<br>(-1.9730400, -0.6397608) | **-0.0013337**<br>(-0.0022172, -0.0004503) | 0.0043738<br>(-0.0481413, 0.0568889) |
| | FVC | **-2.3886740**<br>(-3.025584, -1.7517650) | **-0.0011835**<br>(-0.0021021, -0.0002649) | 0.0032892<br>(-0.0492554, 0.0558337) |

Abbreviations: $FEV_1$, Forced Expiratory Volume in one second; FVC, Forced Volume Capacity, $PM_{2.5}$, (fine) particulates with diameters of 2.5 µm and smaller; BC, black carbon proportion in the fine particulate matter

Table shows the decompositions of the Total Effects of the associations between long-term time-weighted air pollution (in µg/m³) up to baseline and Cognitive Processing Time (CPT) measured by the Cogstate Brief Battery into the single estimation parameters from the used equations. The direct (Direct Effect) and the indirect (Indirect Effect) routes over the lung function measures FEV1 and FVC (potential mediators) at baseline (2006-2012) were modelled simultaneously by performing linear structural equation models with robust standard errors by Huber/ White. Illustrated were the point estimators and confidence intervals coming from single models for each air pollutant, controlled for sex, age, province of residence, educational level, income, pack-years of cigarettes smoked, hypertension, asthma, COPD, diabetes, depression, stroke, multiple sclerosis, BMI, the age- and z-standardized CBB accuracy, and the total number of CBB trials. Shown values for point estimators, lower and upper confidence intervals were not transformed after estimation. Bold values mark significant coefficients.

**Supplementary Table S2.** Sensitivity Analysis of the Model Approach – Causal Mediation Analyses Using G-Formula and Bootstrapping Method

| **Air pollutant** | **Lung function measure** | **Total Effect** | **Indirect Effect** | **Direct Effect** | **Mediation/ Direct Effect** |
|---|---|---|---|---|---|
| | | $\beta_1+\theta_1+\theta_2$ | $\beta_1*\theta_2$ | $\theta_1$ | |
| $PM_{2.5}$ | $FEV_1$ | **18.40** (7.72, 29.28) | **0.32** (0.05, 0.64) | **18.08** (7.34, 29.16) | Mediation + direct effect |
| | FVC | **18.39** (9.00, 27.97) | **0.48** (0.02, 0.96) | **17.92** (8.46, 27.68) | Mediation + direct effect |
| BC | $FEV_1$ | 6.56 (-53.92, 68.09) | **1.73** (0.30, 3.41) | 4.83 (-55.92, 67.50) | Mediation |
| | FVC | 6.55(-54.24, 68.45) | **2.81** (0.08, 5.49) | 3.74 (-57.14, 66.56) | Mediation |

Abbreviations: $PM_{2.5}$, (fine) particulates with diameters of 2.5 µm and smaller; BC, black carbon proportion in the fine particulate matter; $FEV_1$, Forced Expiratory Volume in one second; FVC, Forced Volume Capacity

Table shows the decompositions of the Total Effects of the associations between long-term time-weighted air pollution (in µg/m³) up to baseline and Cognitive Processing Time (CPT) measured by the Cogstate Brief Battery. The direct (Direct Effect) and the indirect (Indirect Effect) routes over the lung function measures $FEV_1$ and FVC (potential mediators) at baseline (2006-2012) were modelled simultaneously by performing g-formula method using bootstrapped standard errors (1,000 iterations). Illustrated were the point estimators and confidence intervals coming from single models for each air pollutant, controlled for sex, age, province of residence, educational level, income, pack-years of cigarettes smoked, hypertension, asthma, COPD, diabetes, depression, stroke, multiple sclerosis, BMI, the age- and z-standardized CBB accuracy, and the total number of CBB trials. Shown values for point estimators, lower and upper confidence intervals were not transformed after estimation. Shown values for point estimators, lower and upper confidence intervals were multiplied by $10^3$. Bold values mark significant paths.

**Supplementary Table S3.** Comparison of the Descriptive Statistics among Participants with Complete (n = 31232) and Incomplete Data (n = 110908)

| **Characteristic** | **Up to baseline** | **Baseline (2006-2012)** | **Follow-up (2014-2015)** | **Up to baseline** | **Baseline (2006-2012)** | **Follow-up (2014-2015)** |
|---|---|---|---|---|---|---|
| | **Complete data** | | | **Incomplete data** | | |
| No. of participants | | n = 31232 | | | n = 110,908 | |
| Sex, No. (%) | | | | | | |
| Women | | 18342 (58.73) | | | 64703 (58.34) | |
| Men | | 12890 (41.27) | | | 46205 (41.66) | |
| Age, mean (SD) | | 43.95 (11.02) | | | 50.10 (12.77) | |
| Province of residence, No. (%) | | | | | | |
| Drenthe | | 5076 (16.25) | | | 29963 (27.02) | |
| Friesland | | 15968 (51.13) | | | 43531 (39.25) | |
| Groningen | | 9386 (30.05) | | | 33711 (30.40) | |
| Other | | 802 (2.57) | | | 3703 (3.34) | |
| Educational level, No. (%) | | | | | | |
| No education | | 61 (0.20) | | | 504 (0.46) | |
| Primary education | | 310 (0.99) | | | 2279 (2.06) | |
| Lower or preparatory vocation | | 3130 (10.02) | | | 14299 (12.95) | |
| Junior general secondary education | | 3909 (12.52) | | | 14973 (13.56) | |
| Secondary vocational education | | 9352 (29.94) | | | 32926 (29.82) | |
| Senior general secondary education | | 3022 (9.68) | | | 9523 (8.62) | |
| Higher vocational education | | 8614 (27.58) | | | 26946 (24.40) | |
| University education | | 2321 (7.43) | | | 6878 (6.23) | |
| Other | | 513 (1.64) | | | 2088 (1.89) | |
| Missing values, No. (% of all participants) | | | | | 492 (0.44) | |
| Net income per month, No. (%) | | | | | | |
| Lower than 1500 Euro | | 4283 (13.71) | | | 15138 (14.07) | |
| 1500 to 2500 Euro | | 8474 (27.13) | | | 28637 (26.61) | |
| Higher than 2500 Euro | | 14688 (47.03) | | | 49010 (45.54) | |
| Do not know/ do not want to tell | | 3787 (12.13) | | | 14838 (13.79) | |
| Missing values, No. (% of all participants) | | | | | 3285 (2.96) | |
| Pack-years of cigarettes smoked, No. (%)[a] | | | | | | |
| Never smokers | | 14895 (47.69) | | | 51304 (48.82) | |
| Lower/ equal than the 50th percentile of ever smokers | | 8482 (27.16) | | | 27311 (25.99) | |

| | | | | |
|---|---|---|---|---|
| Higher than the 50th percentile of ever smokers | | 7855 (25.15) | | 26588 (25.30) |
| Missing values, No. (% of all participants) | | | | 5814 (5.24) |
| BMI, No. (%) | | | | |
| Lower than 25 | | 15026 (48.11) | | 50021 (45.10) |
| 25 to lower than 30 | | 12254 (39.24) | | 44358 (40.00) |
| Higher/ equal than 30 | | 3952 (12.65) | | 16529 (14.90) |
| Hypertension, No. (%)[b] | | | | |
| Yes | | 4869 (15.59) | | 20320 (18.32) |
| No | | 26363 (84.41) | | 90588 (81.68) |
| Asthma, No. (%)[c] | | | | |
| Yes | | 2322 (7.43) | | 8461 (7.63) |
| No | | 28910 (92.57) | | 102447 (92.37) |
| COPD, No. (%)[c] | | | | |
| Yes | | 1342 (4.30) | | 5584 (5.03) |
| No | | 29890 (95.70) | | 105324 (94.97) |
| Diabetes, No. (%)[c] | | | | |
| Yes | | 2056 (6.58) | | 8446 (7.62) |
| No | | 29176 (93.42) | | 102462 (92.38) |
| Depression, No. (%)[c] | | | | |
| Yes | | 3158 (10.11) | | 10455 (9.43) |
| No | | 28074 (89.89) | | 100453 (90.57) |
| Stroke, No. (%)[c] | | | | |
| Yes | | 139 (0.45) | | 787 (0.71) |
| No | | 31093 (99.55) | | 110121 (99.29) |
| Multiple Sclerosis, No. (%)[c] | | | | |
| Yes | | 62 (0.20) | | 255 (0.23) |
| No | | 31170 (99.80) | | 110653 (99.77) |
| Long-term air pollution, mean (SD)[d] | | | | |
| PM2.5 (in µg/ $m^3$) from ELAPSE | 14.95 (0.95) | | 14.99 (2.73) | |
| BC (in µg/ $m^3$) from ELAPSE | 1.25 (0.21) | | 1.24 (0.24) | |
| Total exposure time (in days) for ELAPSE | 8567.56 (2448.31) | | 8974.04 (2603.82) | |
| Missing values, No. (% of all participants) | | | 59 (0.00) | |
| Lung function, mean (SD)[e] | | | | |
| FEV1 (L), % predicted | | 96.62 (12.59) | | 96.33 (13.29) |
| FVC (L), % predicted | | 100.69 (12.07) | | 100.65 (12.67) |

| | | |
|---|---|---|
| Missing values, No. (% of all participants) | | 27620 (24.90) |
| Number of CBB trials, mean (SD)[f] | 139.87 (24.35) | 140.58 (25.01) |
| Missing values, No. (% of all participants) | | 45866 (41.35) |
| Accuracy of given responses, mean (SD)[g] | 0.00 (1.00) | 0.00 (1.00) |
| Less accurate (lower values) than the mean, No. (%) | 14577 (46.67) | 28779 (44.25) |
| More accurate (higher values) than the mean, No. (%) | 16655 (53.33) | 36263 (55.75) |
| Missing values, No. (% of all participants) | | 46240 (41.69) |
| Cognitive processing time (CPT), mean (SD)[h] | 0.00 (1.00) | 0.00 (1.00) |
| Faster CPT (lower values) than the mean, No. (%) | 16103 (51.56) | 31999 (49.20) |
| Slower CPT (higher values) than the mean, No. (%) | 15129 (48.44) | 33043 (50.80) |
| Missing values, No. (% of all participants) | | 46240 (41.69) |

Abbreviations: BMI, body mass index (calculated as weight in kilograms divided by height in meters squared); $PM_{2.5}$, (fine) particulates with diameters of 2.5 µm and smaller; BC, black carbon proportion in the fine particulate matter

[a] Pack-years of cigarettes smoked were calculated from the baseline questionnaire (1 pack-year = 20 cigarettes per day in 1 year).
[b] Hypertension was defined when systolic pressure was higher than 139 mmHg or diastolic pressure higher than 89 mmHg (blood pressure was measured by medical staff).
[c] Prevalence of diseases at baseline was assessed by questions, whether a specific disease was diagnosed by a doctor or not.
[d] Long-term air pollution concentrations were assessed as time-weighted average concentrations up to the 31st December before the baseline year.
[e] Lung function was measured by spirometry by trained medical staff.
[f] Number of CBB trials represents the total number of responses given by the participants during the three subtests (detection, identification, working-memory) of the Cogstate Brief Battery.
[g] Accuracy was measured by the proportion of correct responses. For each domain (detection, identification, working-memory), the accuracy of each response to each trial was recorded. Each of the three already arcsine-transformed and age-standardized accuracy measures from the Cogstate Brief Battery were z-standardized, averaged and again z-standardized.
[h] Cognitive Processing Time was measured by using three (detection, identification, working-memory) log10-transformed and age-standardized speed measures from the Cogstate Brief Battery. The three measures were z-standardized, averaged and again z-standardized.

**Supplementary Table S4.** Descriptive Statistics - Sensitivity Models among Participants Aged 45+ Only and by Using One-Year Annual Concentrations Only

| **Characteristic** | **Up to baseline** | **Baseline (2006-2012)** | **Follow-up (2014-2015)** | **Up to baseline** | **Baseline (2006-2012)** | **Follow-up (2014-2015)** |
|---|---|---|---|---|---|---|
| | **Participants aged 45+ only** | | | **One-Year Annual Concentrations Only** | | |
| No. of participants | n = 15624 | | | n = 49705 | | |
| Sex, No. (%) | | | | | | |
| Women | | 8963 (57.37) | | | 29422 (59.19) | |
| Men | | 6661 (42.63) | | | 20283 (40.81) | |
| Age, mean (SD) | | 52.66 (6.68) | | | 44.77 (11.41) | |
| Province of residence, No. (%) | | | | | | |
| Drenthe | | 2707 (17.33) | | | 10117 (20.35) | |
| Friesland | | 7975 (51.04) | | | 22087 (44.44) | |
| Groningen | | 4603 (29.46) | | | 16402 (33.00) | |
| Other | | 339 (2.17) | | | 1099 (2.21) | |
| Educational level, No. (%) | | | | | | |
| No education | | 42 (0.27) | | | 124 (0.25) | |
| Primary education | | 197 (1.26) | | | 565 (1.14) | |
| Lower or preparatory vocation | | 2131 (13.64) | | | 5549 (11.16) | |
| Junior general secondary education | | 2515 (16.10) | | | 6728 (13.54) | |
| Secondary vocational education | | 4107 (26.29) | | | 15386 (30.95) | |
| Senior general secondary education | | 1419 (9.08) | | | 4538 (9.13) | |
| Higher vocational education | | 3978 (25.46) | | | 12803 (25.76) | |
| University education | | 927 (5.93) | | | 3175 (6.39) | |
| Other | | 308 (1.97) | | | 837 (1.68) | |
| Net income per month, No. (%) | | | | | | |
| Lower than 1500 Euro | | 1577 (10.09) | | | 6497 (13.07) | |
| 1500 to 2500 Euro | | 4368 (27.96) | | | 13413 (26.99) | |
| Higher than 2500 Euro | | 7537 (48.24) | | | 23302 (46.88) | |
| Do not know/ do not want to tell | | 2142 (13.71) | | | 6493 (13.06) | |
| Pack-years of cigarettes smoked, No. (%)[a] | | | | | | |
| Never smokers | | 6204 (39.71) | | | 23773 (47.83) | |
| Lower/ equal than the 50th percentile of ever smokers | | 4272 (27.34) | | | 13184 (26.52) | |
| Higher than the 50th percentile of ever smokers | | 5148 (32.95) | | | 12748 (25.65) | |
| BMI, No. (%) | | | | | | |

| | | | | | | |
|---|---|---|---|---|---|---|
| Lower than 25 | | 6464 (41.37) | | | 22750 (45.77) | |
| 25 to lower than 30 | | 6949 (44.48) | | | 19892 (40.02) | |
| Higher/ equal than 30 | | 2211 (14.15) | | | 7063 (14.21) | |
| Hypertension, No. (%)[b] | | | | | | |
| Yes | | 3375 (21.60) | | | 8597 (17.30) | |
| No | | 12249 (78.40) | | | 41108 (82.70) | |
| Asthma, No. (%)[c] | | | | | | |
| Yes | | 962 (6.16) | | | 3803 (7.65) | |
| No | | 14662 (93.84) | | | 45902 (92.35) | |
| COPD, No. (%)[c] | | | | | | |
| Yes | | 790 (5.06) | | | 2332 (4.69) | |
| No | | 14834 (94.94) | | | 47373 (95.31) | |
| Diabetes, No. (%)[c] | | | | | | |
| Yes | | 1307 (8.37) | | | 3528 (7.10) | |
| No | | 14317 (91.63) | | | 46177 (92.90) | |
| Depression, No. (%)[c] | | | | | | |
| Yes | | 1663 (10.64) | | | 4865 (9.79) | |
| No | | 13961 (89.36) | | | 44840 (90.21) | |
| Stroke, No. (%)[c] | | | | | | |
| Yes | | 112 (0.72) | | | 262 (0.53) | |
| No | | 15512 (99.28) | | | 49443 (99.47) | |
| Multiple Sclerosis, No. (%)[c] | | | | | | |
| Yes | | 32 (0.20) | | | 114 (0.23) | |
| No | | 15592 (99.80) | | | 49591 (99.77) | |
| Long-term air pollution, mean (SD)[d] | | | | | | |
| PM2.5 (in µg/ $m^3$) from ELAPSE | 14.90 (0.98) | | | 14.92 (1.01) | | |
| BC (in µg/ $m^3$) from ELAPSE | 1.23 (0.21) | | | 1.22 (0.22) | | |
| Total exposure time (in days) for ELAPSE | 8655.35 (2455.38) | | | | | |
| Lung function, mean (SD)[e] | | | | | | |
| FEV1 (L), % predicted | | 97.73 (13.59) | | | 96.54 (12.72) | |
| FVC (L), % predicted | | 102.60 (12.75) | | | 100.78 (12.14) | |
| Number of CBB trials, mean (SD)[f] | | | 141.99 (25.50) | | | 140.26 (24.65) |
| Accuracy of given responses, mean (SD)[g] | | | 0.00 (1.00) | | | 0.00 (1.00) |
| Less accurate (lower values) than the mean, No. (%) | | | 6837 (43.76) | | | 22427 (45.12) |
| More accurate (higher values) than the mean, No. (%) | | | 8787 (56.24) | | | 27278 (54.88) |

| | | |
|---|---|---|
| Cognitive processing time (CPT), mean (SD)[h] | 0.00 (1.00) | 0.00 (1.00) |
| Faster CPT (lower values) than the mean, No. (%) | 6837 (43.60) | 24457 (49.20) |
| Slower CPT (higher values) than the mean, No. (%) | 7924 (50.72) | 25248 (50.80) |

Abbreviations: BMI, body mass index (calculated as weight in kilograms divided by height in meters squared); $PM_{2.5}$, (fine) particulates with diameters of 2.5 µm and smaller; BC, black carbon proportion in the fine particulate matter

[a] Pack-years of cigarettes smoked were calculated from the baseline questionnaire (1 pack-year = 20 cigarettes per day in 1 year).
[b] Hypertension was defined when systolic pressure was higher than 139 mmHg or diastolic pressure higher than 89 mmHg (blood pressure was measured by medical staff).
[c] Prevalence of diseases at baseline was assessed by questions, whether a specific disease was diagnosed by a doctor or not.
[d] Long-term air pollution concentrations were assessed as time-weighted average concentrations up to the 31st December before the baseline year.
[e] Lung function was measured by spirometry by trained medical staff.
[f] Number of CBB trials represents the total number of responses given by the participants during the three subtests (detection, identification, working-memory) of the Cogstate Brief Battery.
[g] Accuracy was measured by the proportion of correct responses. For each domain (detection, identification, working-memory), the accuracy of each response to each trial was recorded. Each of the three already arcsine-transformed and age-standardized accuracy measures from the Cogstate Brief Battery were z-standardized, averaged and again z-standardized.
[h] Cognitive Processing Time was measured by using three (detection, identification, working-memory) log10-transformed and age-standardized speed measures from the Cogstate Brief Battery. The three measures were z-standardized, averaged and again z-standardized.

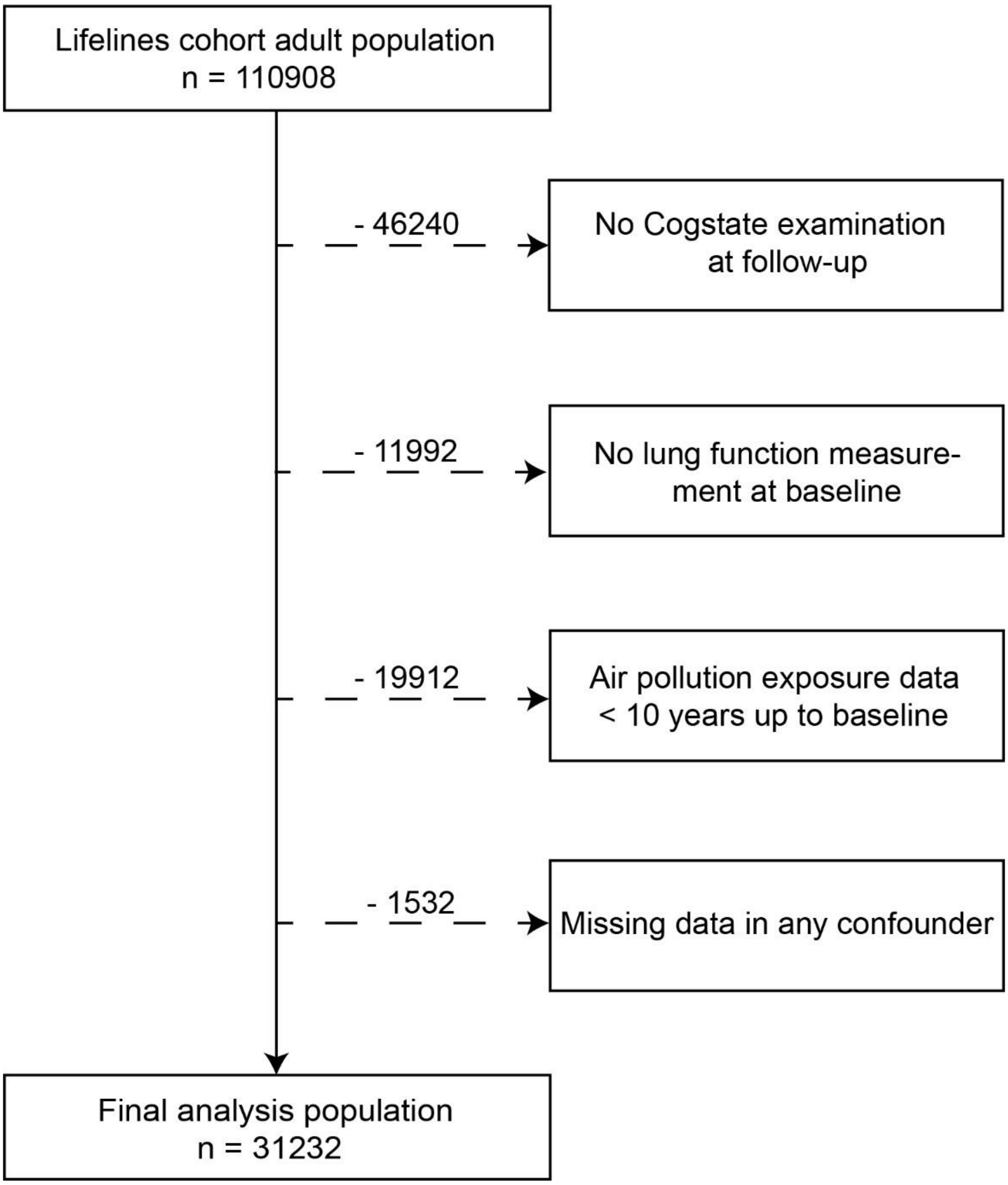


**Supplementary Figure S1.** Construction of the Study Population Based on the Lifelines Cohort Study Population Aged 18+

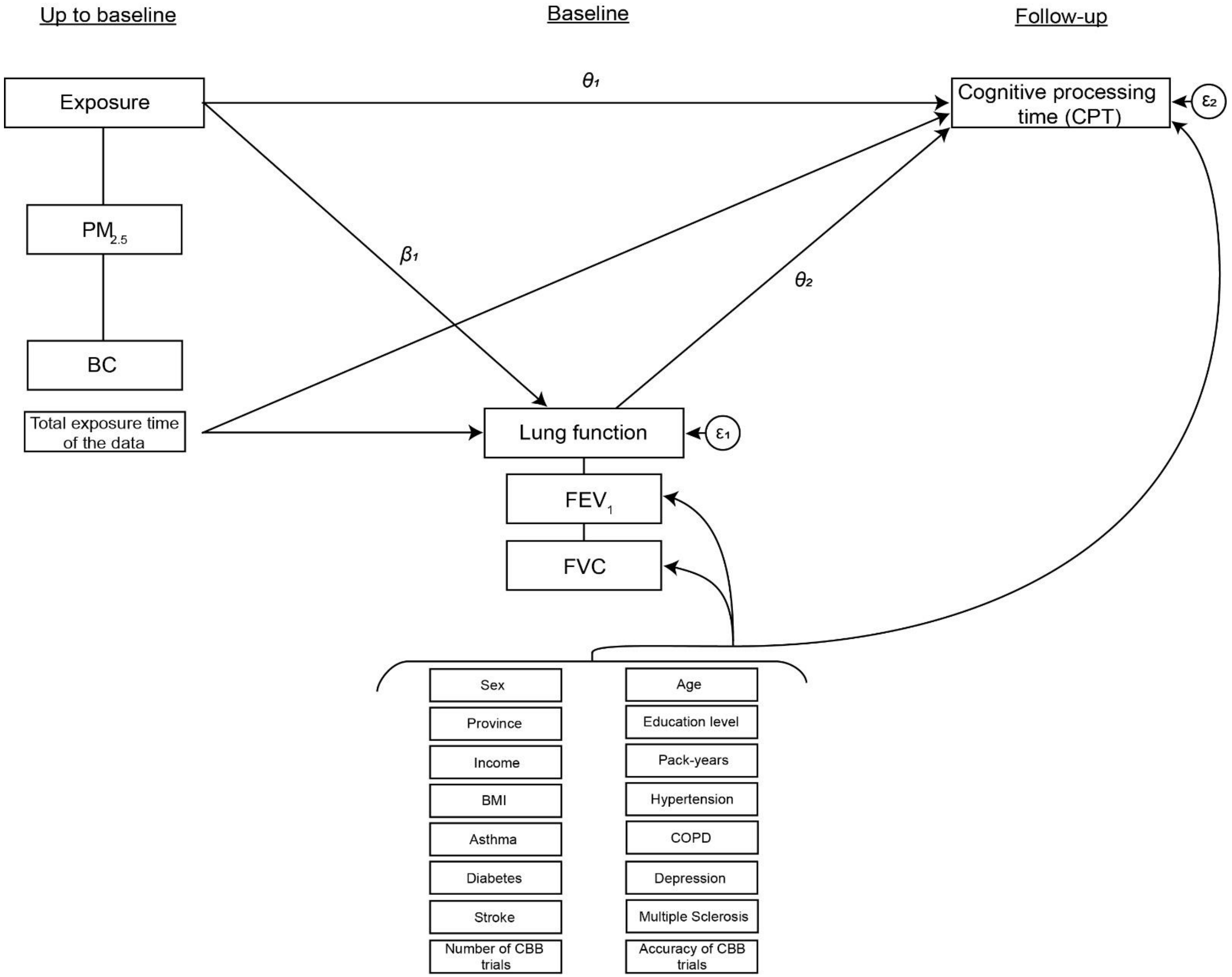


**Supplementary Figure S2.** Used Structural Equation Model (SEM) to Test the Direct and Indirect Routes of Air Pollutants on Cognitive Processing Time (CPT)

**Supplementary Figure S3.** Adjusted Predictions of Cognitive Processing Time for the Total Effects of $PM_{2.5}$

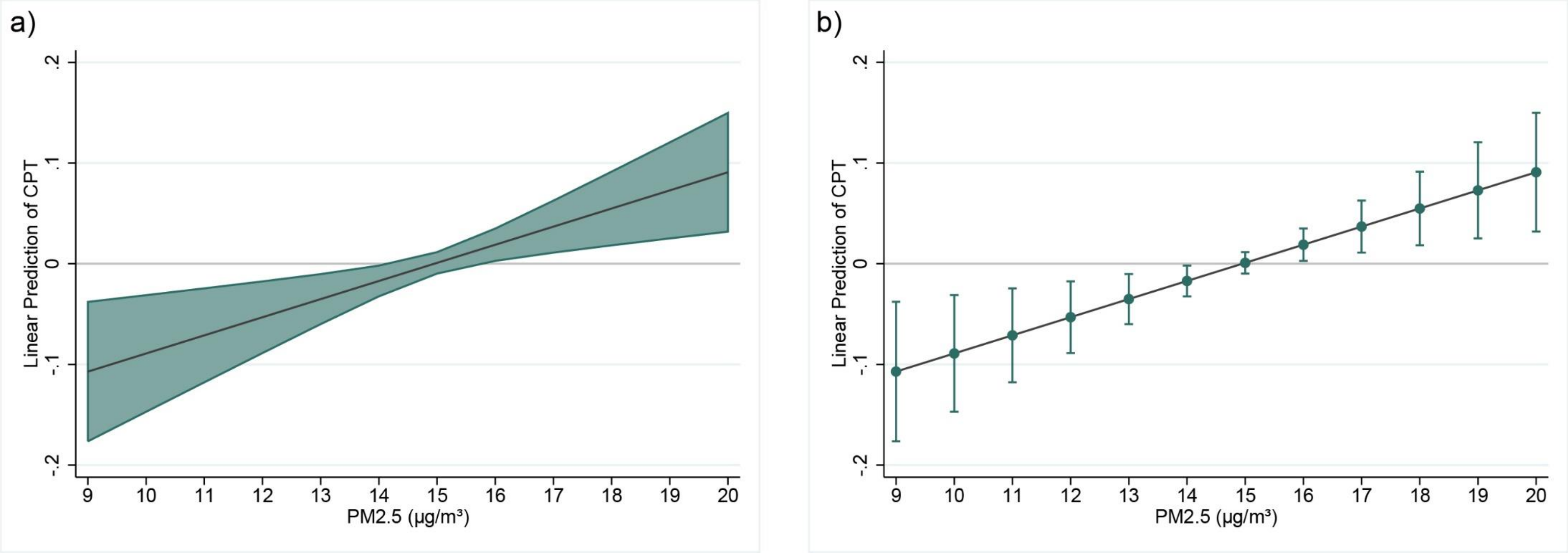


Abbreviations: $PM_{2.5}$, (fine) particulates with diameters of 2.5 µm and smaller; BC, black carbon proportion in the fine particulate matter; CPT, cognitive processing time

Adjusted Predictions of cognitive processing time (CPT) for the Total Effects of $PM_{2.5}$ were estimated for each single integer exposure value between the minimum and the maximum by a linear model. The linear predictions (black line) of CPT were controlled for sex, age, province of residence, educational level, income, pack-years of cigarettes smoked, hypertension, asthma, COPD, diabetes, depression, stroke, multiple sclerosis, BMI, the age- and z-standardized CBB accuracy, and the total number of CBB trials. Green lines show the 95% confidence intervals for the point estimators. The red line marked the upper confidence interval of the prediction when no effect pf $PM_{2.5}$ (zero value of CPT) on CPT is present.

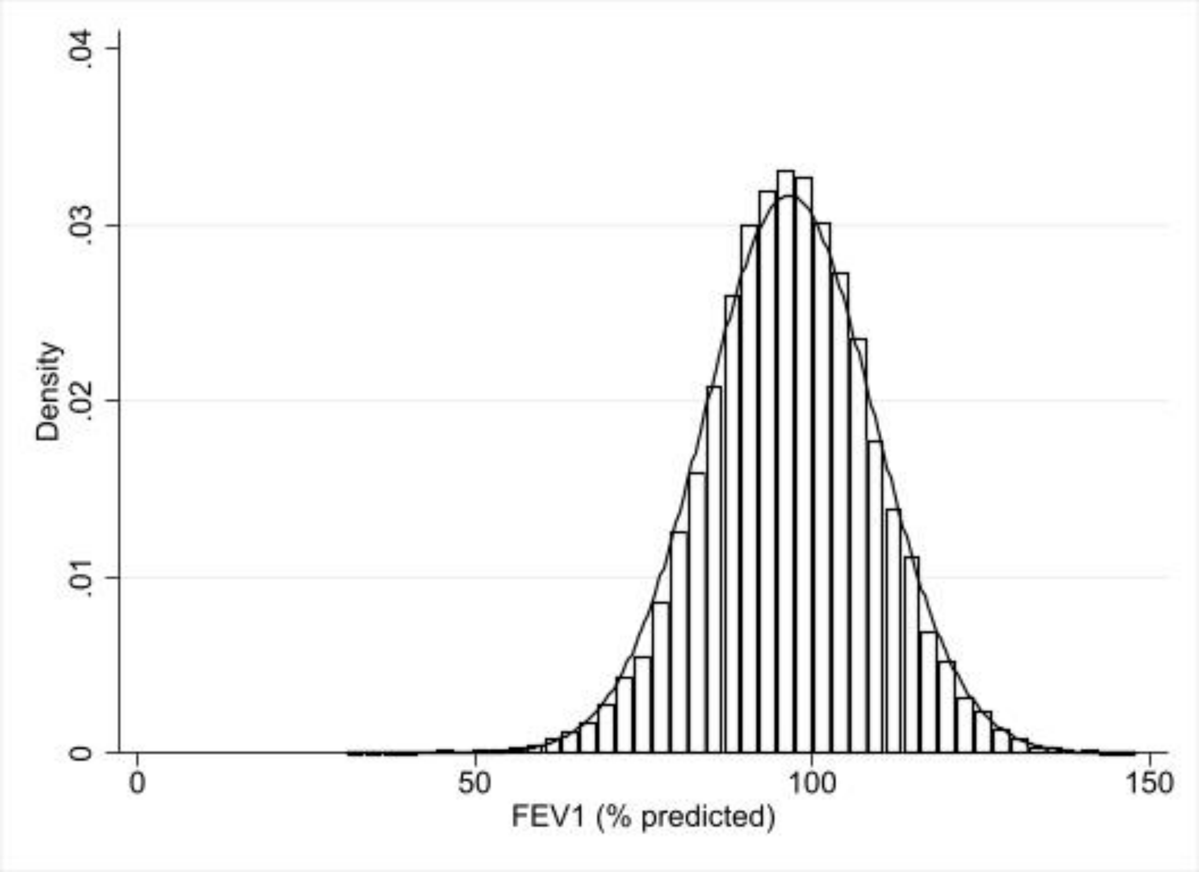


**Supplementary Figure S4.** Distribution of the Mediator Variable ($FEV_1$)

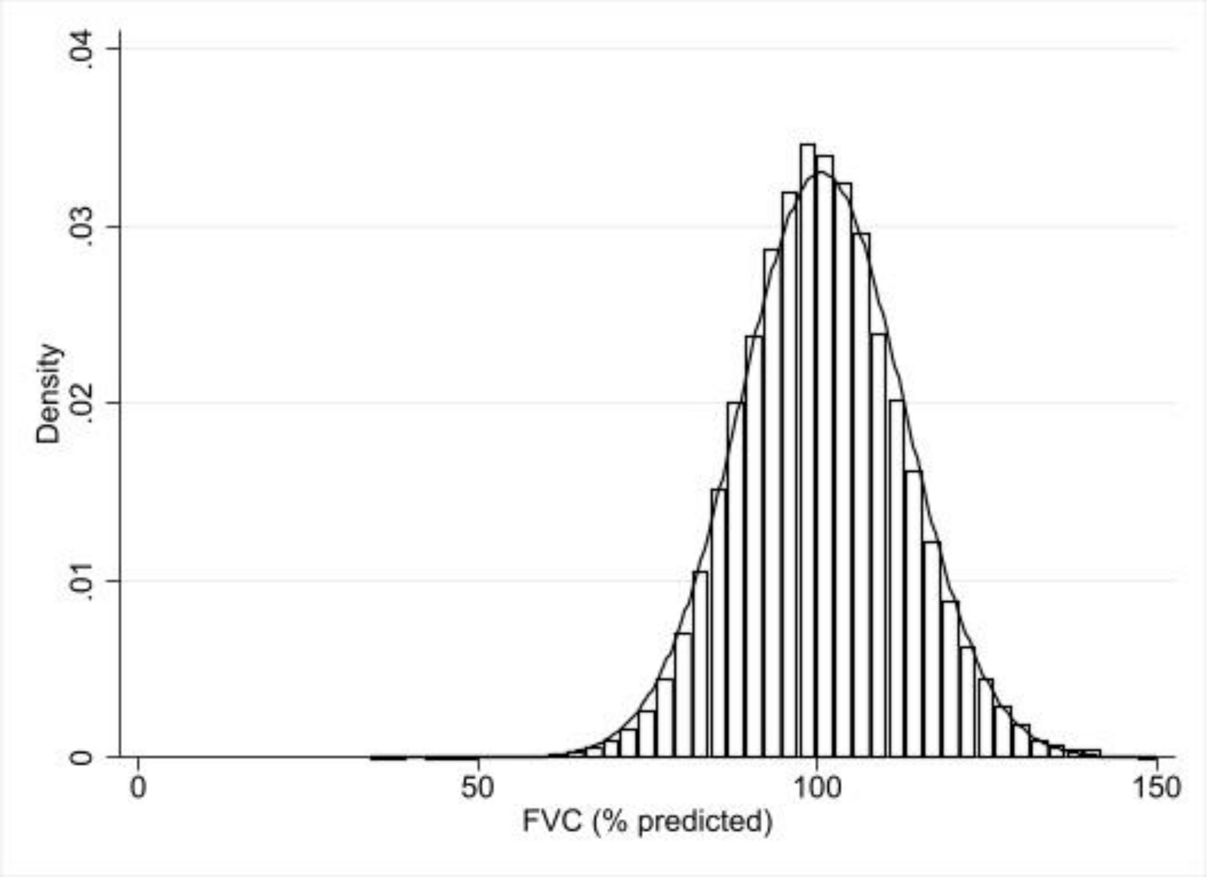


**Supplementary Figure S5.** Distribution of the Mediator Variable (FVC)

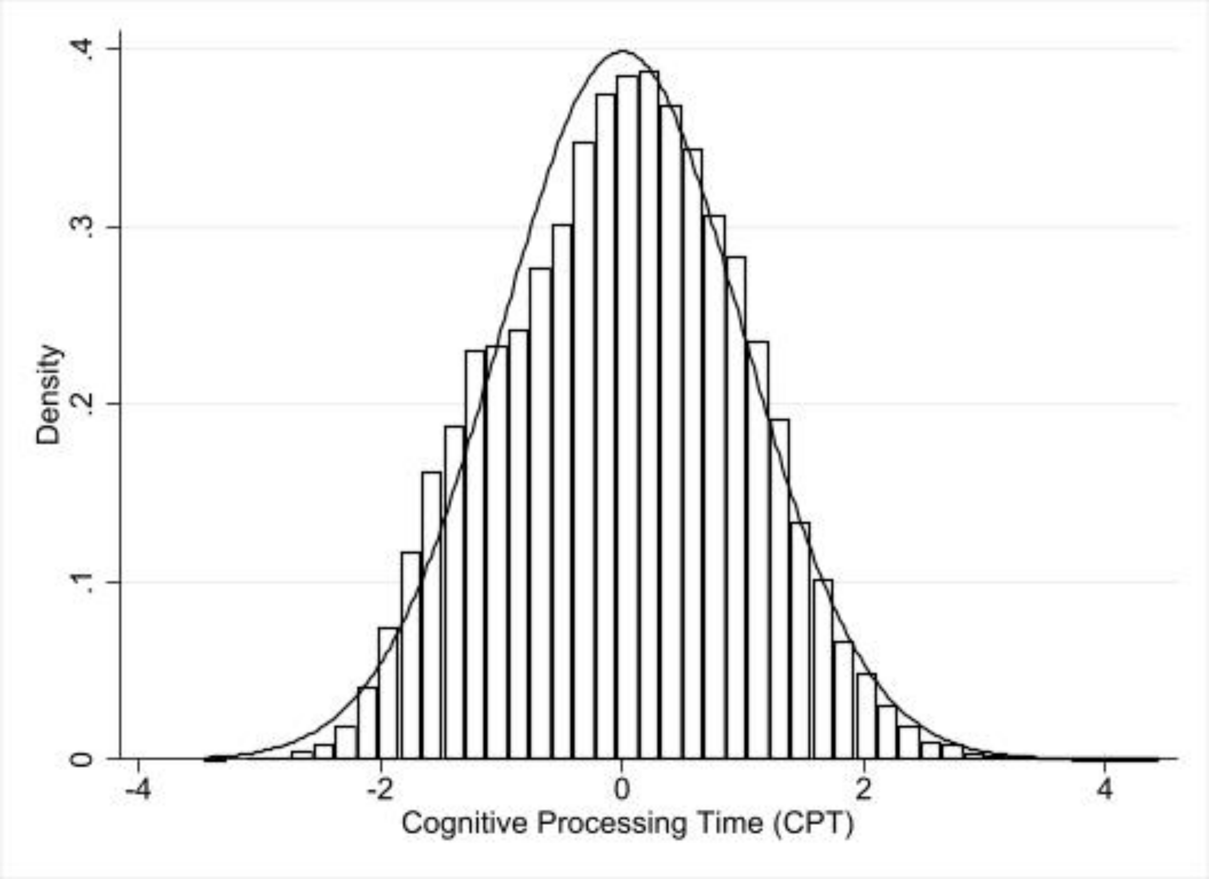


**Supplementary Figure S6.** Distribution of the Outcome Variable (CPT)
